# Infections are not alike: the effects of covariation between individual susceptibility and transmissibility on epidemic dynamics

**DOI:** 10.1101/2024.10.11.24315334

**Authors:** Jeremy D. Harris, Esther Gallmeier, Jonathan Dushoff, Stephen J. Beckett, Joshua S. Weitz

**Author notes:** prior address: Department of Mathematics, Rose-Hulman Institute of Technology, Terre Haute, IN, USA. prior address: School of Biological Sciences, Georgia Institute of Technology, Atlanta, GA, USA. prior address: School of Mathematics, Georgia Institute of Technology, Atlanta, GA, USA. **Corresponding Authors:** Jeremy D. Harris:, Stephen J. Beckett:, Joshua S. Weitz.

## Abstract

Individual-level variation in susceptibility to infection and transmissibility of infection can affect population-level dynamics in epidemic outbreaks. Prior work has incorporated independent variation in susceptibility or transmissibility of individuals into epidemic compartmental models. Here, we develop and assess a mathematical framework that includes covariation in susceptibility and transmissibility. We show that uncorrelated variation in susceptibility and transmissibility leads to an effective transmissibility distribution that has a constant coefficient of variation such that the epidemic dynamics match those with variation in susceptibility alone, providing a baseline for comparison across different correlation structures. Increasing the correlation between susceptibility and transmissibility increases both the speed and strength of the outbreak – and is indicative of outbreaks which might be strongly structured by contact rate variation. In contrast, negative correlations between susceptibility and transmissibility lead to overall weaker outbreaks – with the caveat that the strength of effective transmission increases over time. In either case, correlations can shift the transmissibility distribution, thereby modifying the speed of the epidemic as the susceptible population is depleted. Overall, this work demonstrates how (often unaccounted) covariation in susceptibility and transmission can shape the course of outbreaks and final outbreak sizes.

**Highlights:**

- Developed models incorporating susceptibility and transmissibility covariation.
- Identified eigendistributions of the force of infection.
- Uncorrelated transmissibility reduces to variation in susceptibility alone.
- Positive correlations lead to increases in the basic reproduction number.
- Positive correlations give faster, stronger, more likely outbreaks.
- Effective transmission rates increase over time with negative correlations.

## 1 Introduction

Individuals differ in response to infection: some people may be more likely to get sick than others, and some people may be more likely to transmit an infection on to others. Variation in susceptibility to infection has been introduced into susceptible-infectious-recovered (SIR) (and related) compartmental epidemic models (Kermack & McKendrick, 1927) to account for intrinsic heterogeneity (Rose et al., 2021; Gomes et al., 2022), extrinsic differences based on, e.g., age-dependent contact rates (Davies et al., 2020; Lovell-Read et al., 2022), or differences in prior immunity (Gart, 1972). Prior research has found that variation in susceptibility reduces the epidemic burden (i.e., outbreak size) relative to the homogeneous model (Ball, 1985; Dushoff & Levin, 1995; Coutinho et al., 1999; Dushoff, 1999; Dwyer et al., 2000; Novozhilov, 2008; Novozhilov, 2012; Karev & Novozhilov, 2019; Britton et al., 2020; Rose et al., 2021; Gomes et al., 2022; Tuschhoff & Kennedy, 2024). Hence, the distribution of heterogeneity in susceptibility and the epidemic burden can jointly vary: when susceptible individuals become infected, the joint variation leads to the redistribution and ‘sculpting’ of the susceptibility distribution. The sculpting leads to epidemic slowdowns relative to that of the homogeneous case, reflecting a fundamental difference in the nonlinearity of incident infections. As shown in Rose et al., 2021, the susceptibility distribution is sculpted toward eigendistributions e.g., including gamma distributions with a constant coefficient of variation. Therefore, outbreaks may appear similar during the early stages, but heterogeneity in susceptibility can slow the speed of the epidemic, leading to lower final outbreak sizes (Rose et al., 2021; Gomes et al., 2022), akin to similar impacts of awareness-induced behaviour change (Eksin et al., 2019).

Variability in transmission has also been studied in epidemic models. In practice, the number of secondary infections caused by a focal infected individual can exhibit significant variability, linked to pathogen type, host features, environmental context, and mode of transmission (Lloyd-Smith et al., 2005; Wong & Collins, 2020; Meehan et al., 2023; Murayama et al., 2023). The extent of variation is often measured in terms of an effective ‘dispersion’ parameter of a negative binomial distribution fit to secondary cases (Lloyd-Smith et al., 2005; Lloyd-Smith, 2007; Endo et al., 2020). During the COVID-19 pandemic, household surveys suggest significant variation in both susceptibility and transmissibility (Anderson et al., 2023); and contact survey data has shown that age-dependent variation in contact rates can be a key driver of variation in transmission (Zhang et al., 2020; Quilty et al., 2024) and susceptibility (Britton et al., 2020). Notably, the presence of heterogeneity in transmission may make diseases harder to control at the outset (Frieden & Lee, 2020; Goyal et al., 2022) but can make them more vulnerable to control via mitigation aimed at reducing the relatively small fraction of superspreading events (Frieden & Lee, 2020; Sneppen et al., 2021). Variation both in susceptibility and transmissibility has previously been introduced to epidemic compartmental models. For instance, in a parasite-host system, susceptibility values were fit to dose-response data, revealing that the transmission rate is lower with more heterogeneity (Dwyer et al., 1997). Additionally, models with uncorrelated variation in susceptibility and transmissibility have previously been explored – showing that power-law distributions in transmission can arise from epidemic dynamics unfolding given initial gamma distributed susceptibility and transmissibility (Novozhilov, 2012).

Together, variation in both susceptibility and transmissibility can shape disease dynamics, but compartmental epidemic models have not yet accounted for the effects of potential covariation on dynamics. For example, individuals who interact more with others may be more likely to become infected and more likely to infect others (e.g., if they continue to interact at similar rates when infectious). In this way contact rates could be considered as being equivalent to correlated variation in susceptibility and transmission – but, we caution that many other factors may also be in play. Moreover, individuals may be more vulnerable to infection due to health, behavioural, social, and/or genetic factors that mean they have limited interactions when infectious (e.g., with trained health-care providers) and are therefore less likely to infect others. Here, we advance an approach that is agnostic with respect to contact variation in order to consider both the sign and strength of potential correlations between suceptibility and transmissibility more generally. In this manuscript, we provide a mathematical framework that includes individual-level variation in both susceptibility and transmissibility. The framework allows for comparisons between different model implementations of variation in susceptibility and transmissibility and makes explicit the consequences of covariation between susceptibility and transmissibility on outbreak dynamics.

## 2 Model Framework

### 2.1 Epidemiological dynamics of models with susceptibility and transmissibility

We extend the model framework developed in Rose et al., 2021 to incorporate heterogeneity in both susceptibility and transmissibility into SIR-like epidemic models. To do so, we consider the following population compartment states: susceptible (*S*), infected (*I*), and recovered (*R*). We assume that each individual in the population has a fixed intrinsic susceptibility value, *ε*, as well as fixed intrinsic transmissibility value, *δ*. Hence, the *S*-*I*-*R* compartments are functions of susceptibility (*ε*) and transmissibility (*δ*) that we denote as *S*(*t, ε, δ*), *I*(*t, ε, δ*), and *R*(*t, ε, δ*). We denote *S*(*t*), *I*(*t*), *R*(*t*) to represent the respective population densities of all susceptible, infected and recovered individuals. Then we can define sub-population densities: the susceptible sub-population density with intrinsic susceptibility *ε* and intrinsic transmissibility *δ* is given by

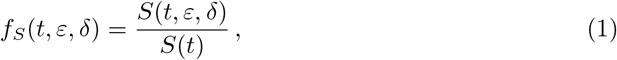

the infected sub-population density is

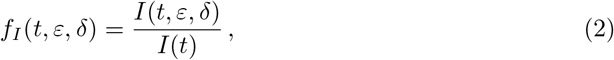

and the recovered sub-population density is

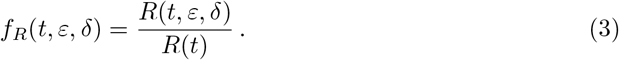

Note that these definitions are joint densities such that at any time, *t*, then 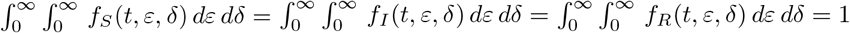 To calculate the mean susceptibility and transmissibility over time, we consider the marginal distributions relevant to the disease dynamics:

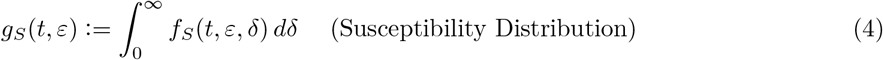

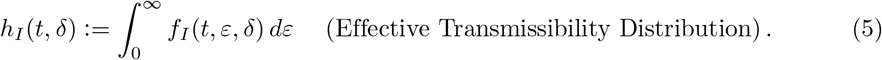

The other relevant marginal distribution is the *potential* transmissibility distribution in the susceptible population. This distribution is indicative of the remaining infectivity of the population who might have the potential to be infectious in the future. During the exponential growth phase of the epidemic individuals are drawn from the susceptible population at varying rates that depend on an individual’s susceptibility to infection. As individuals become infected, their contribution to *effective* transmissibility is drawn from the *potential* transmissibility distribution, and thus, the *effective* transmissibility distribution (Equation 5) is “filled in” by the *potential* transmissibility distribution given by:

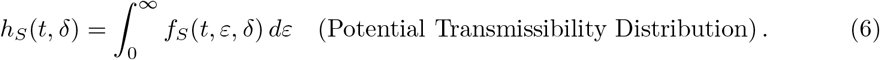

Given the susceptibility distribution (Equation 4) and transmissibility distributions (Equations 5-6), we can define the mean susceptibility and transmissibility. The mean susceptibility to infection within the susceptible population is:

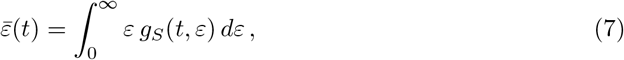

the mean *effective* transmissibility of individuals within the infected population is:

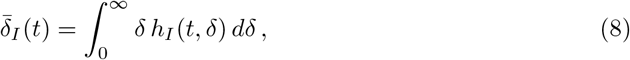

and the mean *potential* transmissibility of individuals in the susceptible population is:

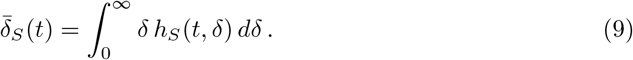

Then the force of infection to the susceptible population with susceptibility level *ε* is:

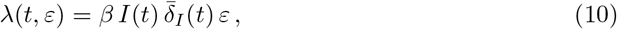

where *β* is the baseline transmission rate. For each subpopulation with (*ε, δ*), the SIR model equations with susceptibility and transmissibility heterogeneity can be written as:

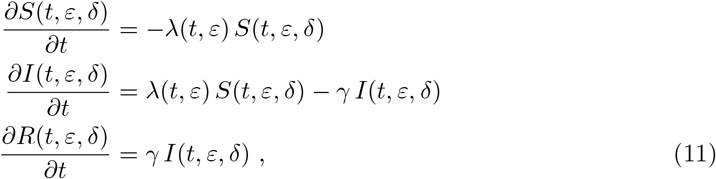

where *γ* is the recovery rate of all infected individuals. See Appendix A for the derivation of Equation 11 from discrete model variables. See Figure 1 for a visual representation of the model framework using discrete susceptibility and transmissibility variables.

**Figure 1.**
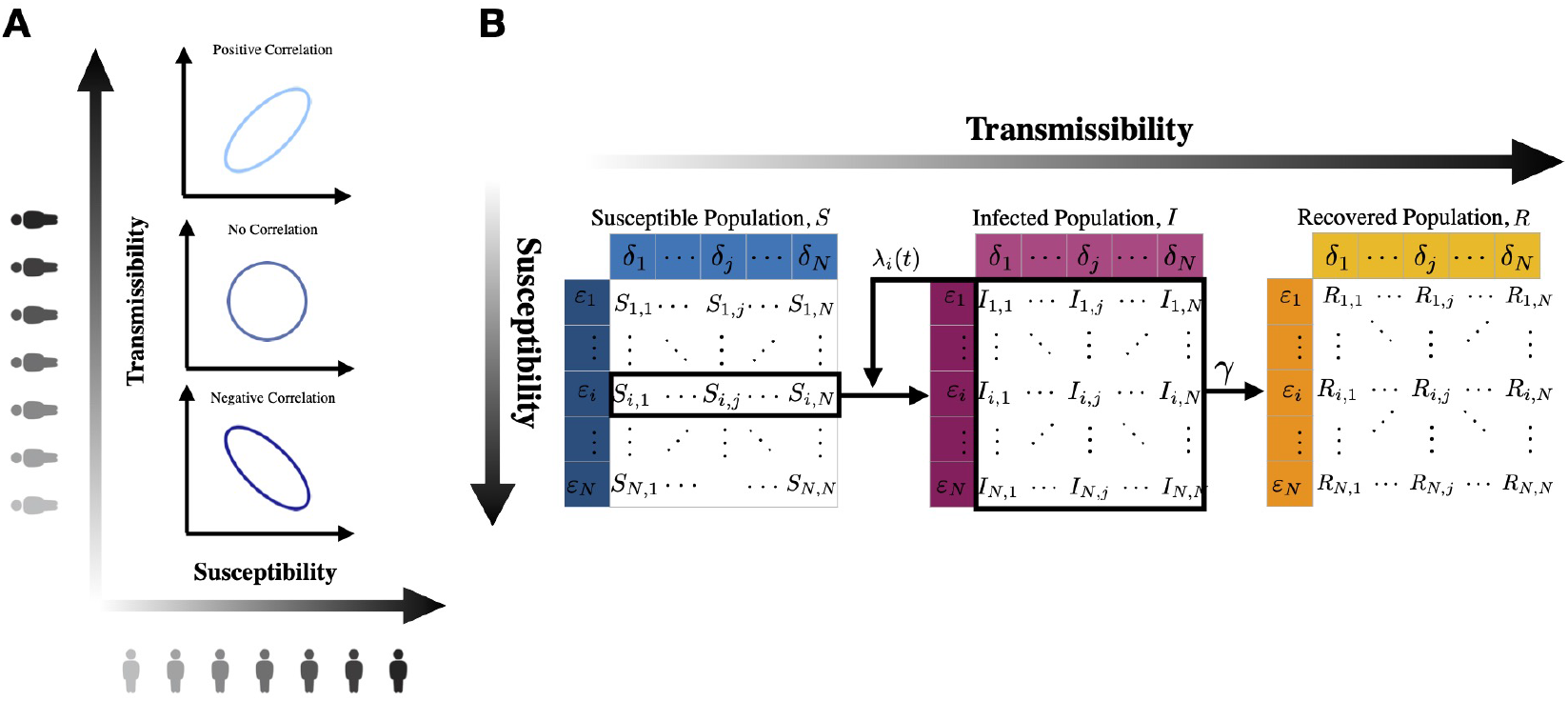
Model diagram: susceptible (*S*), infected (*I*), and recovered (*R*) populations with individual variation in susceptibility and transmissibility. **(A)** Positive correlation, no correlation, and negative correlation between individual susceptibility and transmissibility. **(B)** S-I-R compartments are discretized into subpopulations distributed according to susceptibility (*ε*_*i*_) and transmissibility (*δ*_*j*_). Here, *λ*_*i*_(*t*) = *λ*(*t, ε*_*i*_) is the force of infection to the susceptible population with susceptibility, *ε*_*i*_, and *γ* is the mean recovery rate for all infected individuals.

Integrating with respect to the continuous variables, *ε* and *δ*, we can obtain the total population incidence:

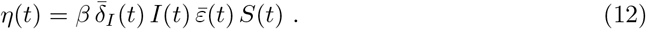

From here, differential equations for the following variables can be identified: the mean susceptibility 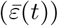 the mean potential transmissibility 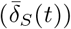 and the mean effective transmissibility 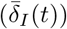. This model can be described by the following 6-dimensional system of ordinary differential equations (where, for convenience, we set *C*_*S*_(*t*) = cov_*S*_(*ε, δ*)(*t*) to represent the covariance between susceptibility and transmissibility in the susceptible subpopulation):

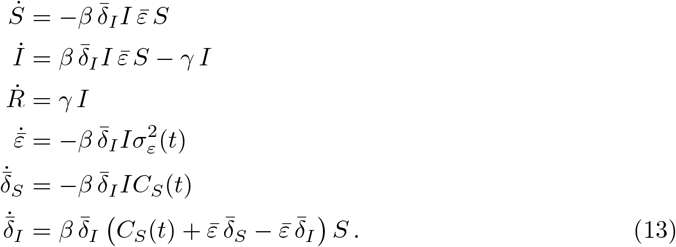

Here, the dependence on *t* is implicit for all time-dependent variables except for the covariance between susceptibility and transmissibility in the susceptible subpopulation, *C*_*S*_(*t*), and the variance, 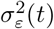, of the susceptibility distribution, *g*_*S*_(*t, ε*). This system is not closed for arbitrary starting joint distributions in the susceptible population, as *C*_*S*_(*t*) and 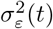 may change over time, impacting the mean susceptibility and transmissibility as the epidemic progresses.

### 2.2 Associations of correlations with epidemic strength and dispersion

Using the dynamical system presented in Equation 13, we can define the basic reproduction number (ℛ_0_) and dispersion (*κ*) of epidemics as a function of the correlation coefficient (*ρ*) between susceptibility and transmissibility. First, in a fully susceptible population, *S* = 1, the basic reproduction number is given by:

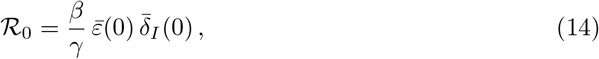

where 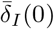 is the mean effective transmissibility during initial exponential growth, and 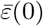 is the initial mean susceptibility. Note that without correlations (i.e., *ρ* = 0) between initial susceptibility and transmissibility values, then 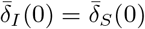, but with correlations (i.e., *ρ ≠* 0), they can differ. During exponential growth, the joint distribution of the susceptible population is fixed, which implies that the derivatives, 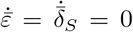. Hence, both *ε* and 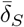 remain fixed during the exponential growth phase. Since the covariance of the joint distribution of the susceptible population, *C*_*S*_(*t*), also remains fixed during the exponential growth phase, then 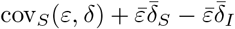 is constant. Hence, in order to obtain the reproduction number, we need to find *δ*_*I*_ (0) relative to *δ*_*S*_(0). In order for 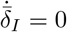 during exponential growth, then 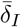 must converge to 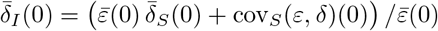, and the reproduction number is

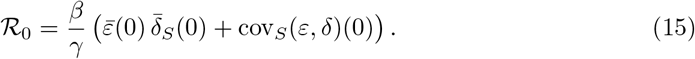

Then the reproduction number can be written equivalently as:

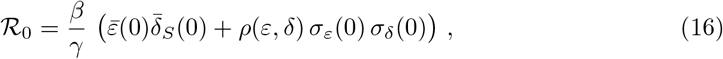

where the correlation coefficient between susceptibility and transmissibility (during exponential growth) is given by 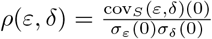.

Second, to define the dispersion, *κ*(*t*), of an epidemic we first let the mean susceptibility, 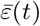, serve as a dimensionless progress variable. Rose et al., 2021 used 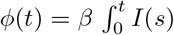 *ds* as a dimensionless progress variable, noting that *ϕ*(*t*) is proportional to the cumulative infectious force. We compute the mean susceptibility 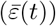, potential transmissibility 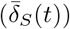, and effective transmissibility 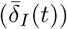 as the epidemic progresses over time. We also compute the variances 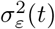 and 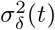 in order to obtain the corresponding squared coefficients of variation over time.

From Equation 13, we follow the analysis in Rose et al., 2021 and have that 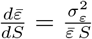. Using differential notation, *d*(ln(*x*)) = *dx/x*, then the square of the coefficient of variation for the susceptibility distribution (*CV* ^2^) is given by:

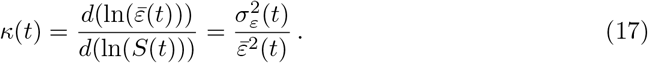

For given initial distributions, we compute the squared coefficient of variation for the susceptibility distribution as well as the squared coefficients of variation for the potential and effective transmissibility distributions over time.

## 3 Results

### 3.1 Uncorrelated gamma-distributed susceptibility and transmissibility

We first examine the dynamics of Equation 13 when susceptibility and transmissibility are uncorrelated. In this case, the covariance, *C*_*S*_, is zero. Hence, from Equation 13, 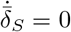 and so 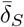 remains constant over time. If the initial potential and effective transmissibility values are equal, i.e., 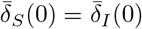, then 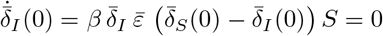. Since 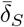 is constant, then 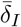 remains constant and equal to the initial potential value, 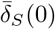. Therefore, without correlations, Equation 13 further simplifies to

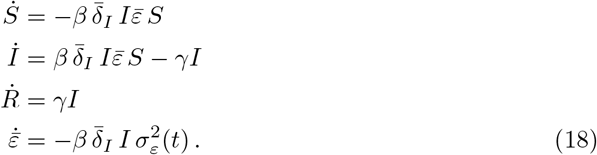

This nonlinear dynamical system is equivalent to prior work on variation in susceptibility alone (Rose et al., 2021; Gomes et al., 2022). Here, the variance in susceptibility 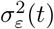 denoted with explicit time *t* to emphasize that it may change over the course of the outbreak. In this work, we also introduce a variation of Equation 18, which we term the ‘reduced model’, in which we approximate the variance term 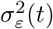 using analytical results from (Rose et al., 2021). For uncorrelated gamma distributions, the reduced model sets the variance 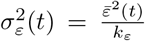 where *k*_*ε*_ is the shape parameter of the gamma distributed susceptibility distribution *g*_*S*_(*ε, t*). For uncorrelated Gaussian distributions, the reduced model sets the variance 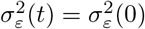.

To examine how the initial distributions changes through the epidemic dynamics, we first consider initially gamma-distributed susceptibility and transmissibility values and examine the squared coefficient of variation in susceptibility, 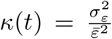 (see Equation 17). For initially uncorrelated gamma distributions, we find that: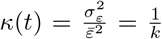, where *k* is the shape parameter of the susceptibility distribution, *g*_*S*_(0, *ε*). Here, the mean transmissibility, 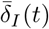, is a multiplicative factor that modifies the force of infection. However, since 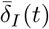 is constant, the system collapses to the system with variation in susceptibility alone. Hence, initially gamma-distributed susceptibility distributions remain gamma-distributed with mean: 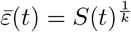 (Rose et al., 2021; Gomes et al., 2022). That is, the same power-law relationship between susceptibility (*ε*) and the susceptible population (*S*(*t*)) holds here with uncorrelated gamma-distributed variation in susceptibility and transmissibility.

We can analyze the change in the joint distribution of susceptibility and transmissibility in the susceptible population, *f*_*S*_(*t, ε, δ*), through the epidemic dynamics. We find that *f*_*S*_(*t, ε, δ*) satisfies the partial differential equation:

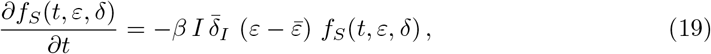

see Appendix B.1. Note that we can integrate Equation 19 over all transmissibility values so that the marginal susceptibility distribution *g*_*S*_(*t, ε*) satisfies:

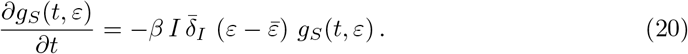

It has been shown (see Section S3 in Rose et al., 2021) that distributions of the exponential family, including initially uncorrelated gamma distributions, with shape parameter *k*, of the form:

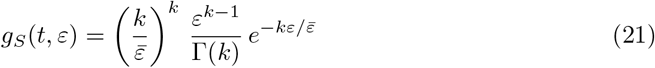

satisfy the PDE given in Equation 20. We show that gamma distributions that evolve with a fixed shape parameter and Gaussian distributions that evolve with a fixed variance satisfy Equation 20 (see Appendix B.2).

To verify this analysis, we can compare the simulations of the discrete model given in Equation 11 with the uncorrelated reduced model in Equation 18, which sets the variance 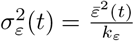 Details of model parameterization and simulation are given in Appendix C-E.

For initially uncorrelated gamma-distributed susceptibility and transmissibility, the dynamics of incident infections (Figure 2A, blue) agree with Equation 18 (Figure 2A, green dashed). They also agree with the case of variation in susceptibility alone (Figure 2A, dashed black). Consistent with results of Rose et al., 2021, variation in susceptibility slows down incident infections compared to the conventional SIR model (Figure 2A, gray). As predicted, the *κ* for both susceptibility transmissibility remain constant over time (Figure 2B,C). We show the initial joint distribution, *f*_*S*_(0, *ε, δ*), in the susceptible population with uncorrelated susceptibility (*ε*; *x*-axis) and potential transmissibility (*δ*; *y*-axis) (Figure 2D). In Figure 2E, we compare the susceptibility distribution, *g*_*S*_(*t, ε*), at two time points during exponential growth: *t*_0_ = 0 days (black circle) and *t*_1_ = 10 days (violet circle). The distribution remains constant during exponential growth when susceptible depletion is negligible. In Figure 2F, we show the potential and effective transmissibility distributions at these time points. The epidemic is initialized with a few infected individuals, and the effective transmissibility distribution, *h*_*I*_ (*t, δ*), in the infected population is determined by potential transmissibility, *h*_*S*_(*t, δ*), in the susceptible population. Thus, to remove transients, we make the initial transmissibility distributions equal: *h*_*I*_ (0, *δ*) = *h*_*S*_(0, *δ*). Then, we show that *h*_*I*_ (0, *δ*) = *h*_*I*_ (*t*_1_, *δ*) remains fixed during exponential growth (Figure 2F, gray matches dashed violet). These results indicate that initially uncorrelated gamma distributions remain gamma-distributed such that the mean susceptibility satisfies 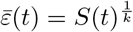, and the effective mean transmissibility 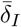 remains constant such that the transmissibility distribution *h*_*I*_ (*t, δ*) is constant over the course of the epidemic.

**Figure 2.**
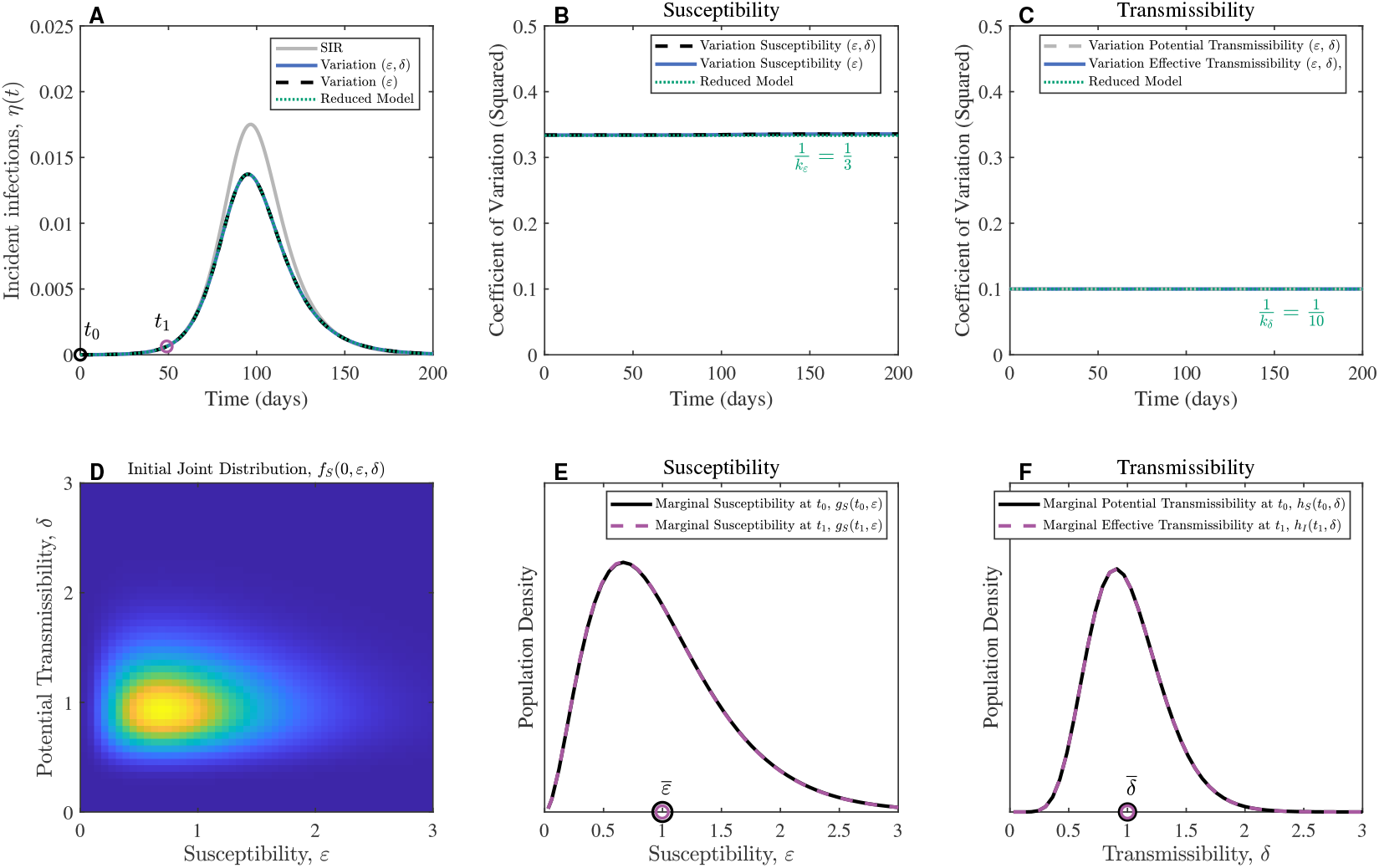
Uncorrelated gamma distributions for susceptibility and transmissibility distributions during exponential growth. **(A)** Incident infections. **(B)** The coefficient of variation (squared) for susceptibility remains constant over time for gamma-distributed susceptibility values. **(C)** The coefficient of variation (squared) in transmissibility remains constant over time for gamma-distributed transmissibility values. **(D)** Initial joint distribution (uncorrelated gamma distributions) for susceptibility values (*ε*) and potential transmissibility values (*δ*). **(E)** Comparing susceptibility distributions at time points: *t*_0_ = 0 and *t*_1_ = 50 days. **(F)** Potential and effective transmissibility distributions at time points: *t*_0_ = 0 and *t*_1_ = 50 days. The transmission rate is equal to *β* = 0.2, and the recovery rate is equal to *γ* = 1*/*10 such that the basic reproduction number is ℛ_0_ = 2.0. Initial gamma distribution shape parameters: *k*_*ε*_ = 3, *k*_*δ*_ = 10. The reduced model refers to Equation 18 with 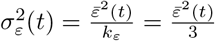.

### 3.2 Uncorrelated Gaussian-distributed susceptibility and transmissibility

As a prelude to introducing correlations we consider initially uncorrelated truncated Gaussian distributions with low (Figure S1) and high (Figure 3) variance, such that *ε, δ >* 0. For the low variance case we show incident infections from discrete model simulations (using Equation 11) in Figure S1A. The coefficient of variation (squared) for susceptibility is constant during the initial exponential growth phase of the outbreak, but increases during susceptible depletion because the mean decreases faster than the variance (see Figure S1B). Here, we set *σ*_*ε*_(*t*) = *σ*_*ε*_(0) in simulations of the reduced model (using Equation 18) and observe increases in *κ*(*t*) (Figure S1B), meaning that the mean susceptibility decreases. As predicted by our analysis, in reducing Equation 13 in the absence of covariation to Equation 18, the coefficient of variation (squared) for transmissibility remains constant over time (Figure S1C). Despite the differences in *κ* of susceptibility between the full and reduced models, the reduced model can still approximate incident infections. Consistent with results from Rose et al., 2021, Gaussian and gamma distributions are eigendistributions with respect to the epidemic dynamics. Moreover, gamma distributions have constant *κ*, whereas Gaussian distributions have constant variance (approximately, considering that Gaussian distributions have proper support on the whole real line).

**Figure 3.**
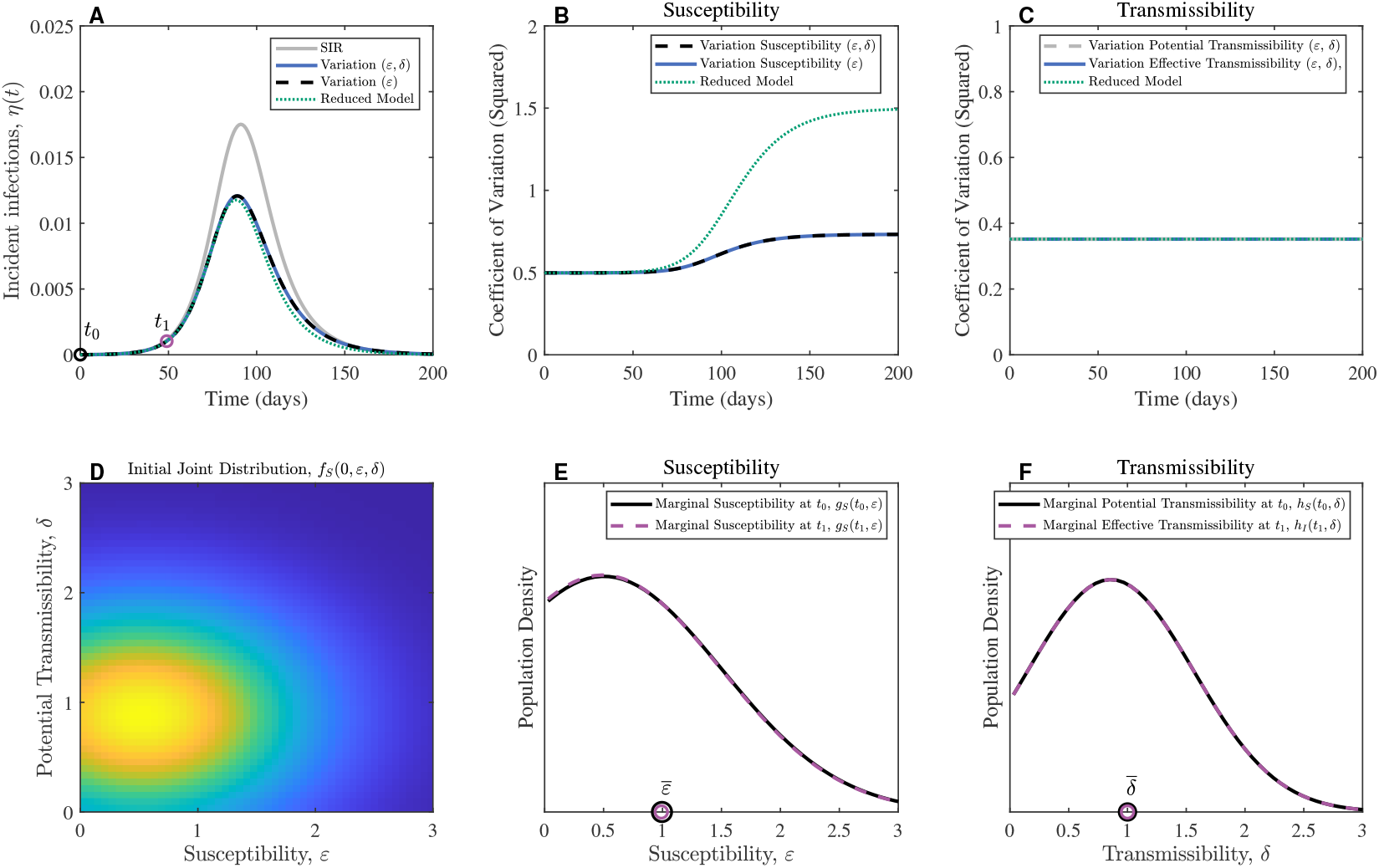
Uncorrelated (high variance) Gaussian distributions for susceptibility and transmissibility during exponential growth. Similar to Figure S1 but with higher variances in the initial susceptibility and transmissibility distribution. **(A)** Incident infections. **(B)** *κ* for susceptibility. **(C)** *κ* for transmissibility. **(D)** Initial joint distribution of susceptibility and transmissibility in the susceptible population, *f*_*S*_(*t*_0_, *ε, δ*), is given by uncorrelated Gaussian distribution, truncated to have positive support in both *ε* and *δ*. **(E)** Susceptibility distributions at *t*_0_ and *t*_1_. **(F)** Potential transmissibility distribution at *t*_0_ matches the Effective transmissibility distribution during exponential growth at *t*_1_. Parameters: The transmission rate is *β* = 0.2, and the recovery rate is *γ* = 1*/*10 such that the basic reproduction number is ℛ_0_ = 2.0. The variance values in the initial joint: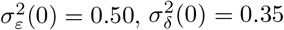, The reduced model refers to Equation 18 with 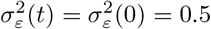.

To better see the effects of correlations when incorporated, we also examine a high variance case in which we increase the variance for both the initial potential susceptibility and transmissibility distributions 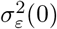 from 0.15 to 0.5; and 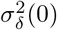 from 0.05 to 0.35). In doing so, the effects of truncation on the bivariate distribution become more pronounced. Even when the initial variances are increased, we can see reasonable agreement, albeit less than with smaller variance, between the full and reduced model simulations (Figure 3A). For truncated Gaussian initial distributions, the *κ* in the reduced model simulations increase more than in the discrete model simulations (Figure 3B). Due to truncation, susceptibility variance decreases over time as well as the mean susceptibility. The reduced model *κ* diverges as this model is unable to capture this decrease in variance over time, as by definition 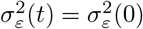 Despite this, the reduced model in Equation 18 provides a reasonable approximation of incident infections in the discrete model simulations with the highest discrepancies observed during the decay phase (Figure 3A).

### 3.3 Correlations between susceptibility and transmissibility modify the basic reproduction number

Next, we introduce covariation by considering correlations between susceptibility and transmissibility and compare the ensuing epidemic dynamics with respect to the case without correlations. We vary the correlation coefficient (*ρ*) from negative to positive, in simulations we explore scenarios over the range of values from −0.6 to 0.6, and find that the speed, i.e., the exponential growth rate, increases with increasing correlation (Figure 4A). Recall that the basic reproductive number is dependent on the correlations between susceptibility and transmissibility (Equation 16). In the absence of correlations (*ρ* = 0), the basic reproduction number is 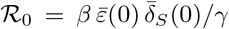, and the product of the initial mean susceptibility and transmissibility multiply the basic reproduction of the conventional SIR model, *β/γ*. Note that ℛ_0_ is an increasing function of the correlation coefficient, *ρ*, which is in agreement with simulations (Figure 4B). For *ρ >* 0, the more susceptible individuals are infected earlier and are also more transmissible than on average, causing more transmission during exponential growth such that the basic reproduction number is greater than in the uncorrelated case. For *ρ <* 0, the basic reproduction number is less than in the uncorrelated case because the more susceptible individuals are less transmissible on average (Figure 4B). For a given transmission rate, *β*, and recovery rate, *γ*, the initial speed and strength of the epidemic increases with the initial correlation coefficient between susceptibility and transmissibility, leading to larger outbreaks by larger initial correlation coefficients (Figure 4A).

**Figure 4.**
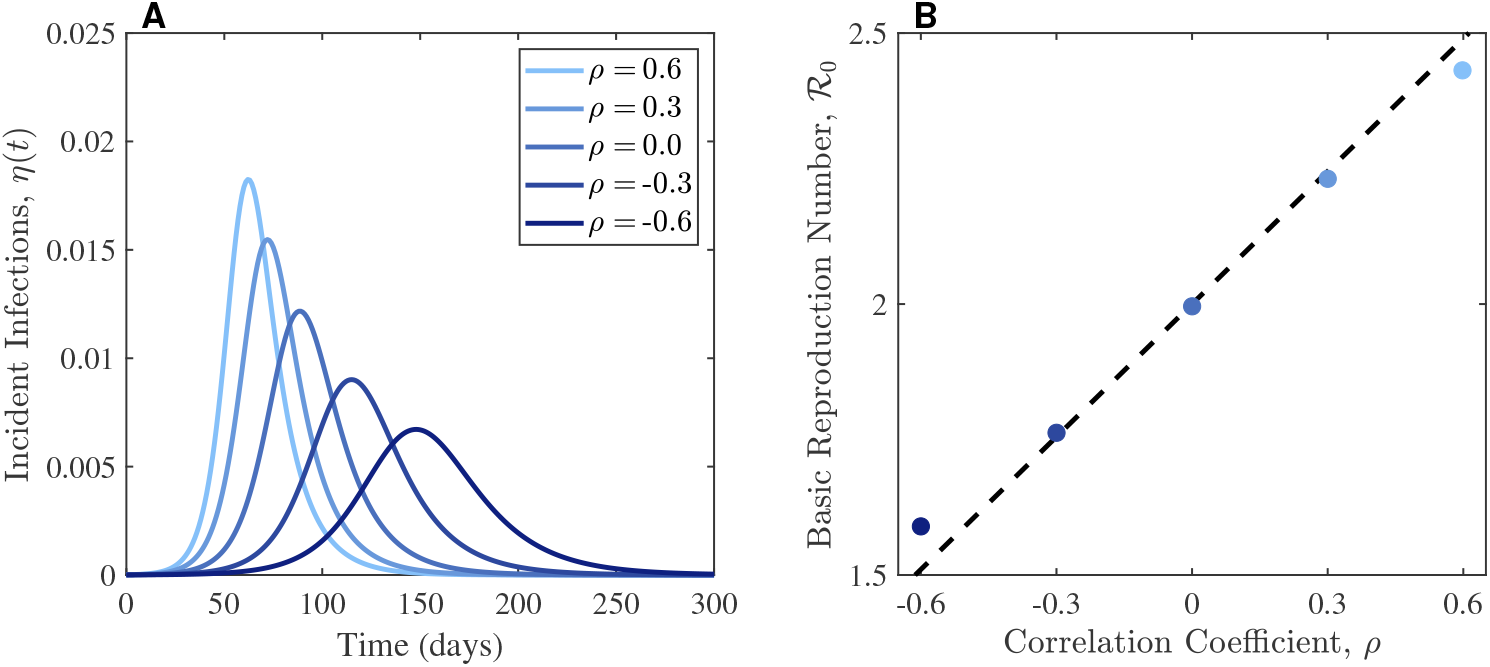
The effects of correlations on the speed and strength of the epidemic. **(A)** The speed of incident infections is the exponential growth rate which varies with the initial correlation coefficient. Positive correlations between susceptibility and transmissibility result in faster epidemic speeds with increased peak incident infections. **(B)** The basic reproduction increases with increasing initial correlation coefficient. Comparison of Equation 14 with computed 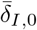, for the five simulations (colored dots corresponding to the scenarios in **A**) and Equation 16 with (approximately) fixed initial standard deviations of the initial joint susceptibility distribution (dashed line). Across all simulations, the means of the initial joint in susceptibility (i.e., *ε*_*S*_(0) = *δ*_*S*_(0) = 1), the transmission rate is equal to *β* = 0.2, and the recovery rate is equal to *γ* = 1*/*10 so that when there is no correlation between susceptibility and transmissibility (*ρ* = 0), then ℛ_0_ = 2, as expected in the standard SIR framework. Varying *ρ* = 0.6, ™0.30.3, 0, 0.3, 0.6, the parameter values of initial joint distribution are given by: 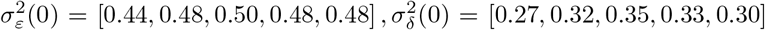 See Figure S2 for the corresponding initial joint distributions.

### 3.4 Sensitivity of heterogeneous model outcomes to an outbreak index case

The introduction of population variability also raises questions about how the initiation of an outbreak may effect epidemic outcomes – the transmissibility and susceptibility of individuals in the the first chains of infection may have a large effect on how an outbreak takes off (see Goyal et al., 2022). To probe this, we first assess how variation in the initial distribution of the infected population may impact epidemic trajectory and timing; and second, utilize stochastic simulations to additionally assess variation in outbreak occurrence and epidemic trajectories. In Figure 5A, we show two potential points in susceptibility-transmissibility parameter space that an index infection could take, and examine the impact of these initial conditions on epidemic trajectories in Figure 5B. We find that differences in the transmissibility (but, not susceptibility) of the initial infection can impact the timing of the epidemic – essentially translating the epidemic trajectory in the time axis. When more infectious individuals seed an outbreak, the epidemic trajectory emerges earlier than if the initial case is less infectious than average – in which case the epidemic occurs more slowly than expected under the outbreak eigendistribution and baseline SIR models. As the infection has already occurred, the susceptibility of this (small) index infection does not play a role in ongoing transmission or the long-term epidemic trajectory given mass-action kinetics.

**Figure 5.**
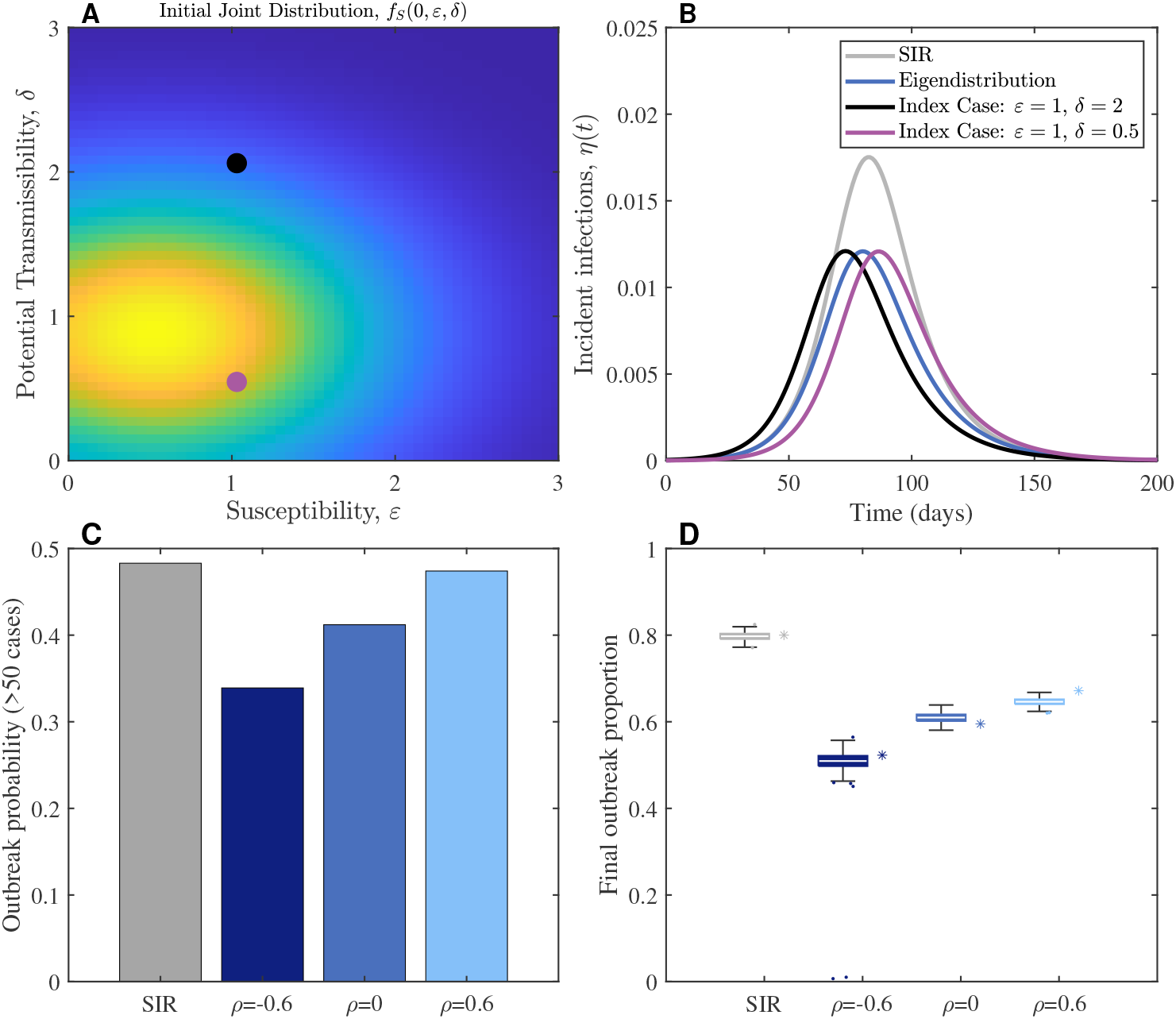
Sensitivity of heterogeneous models with respect to an outbreaks index case. **(A)** Two different choices for the characteristics of the initial index case index with susceptibility *ε* = 1 and transmissibility values: *δ* = 0.5 (gray dot) and *δ* = 2 (black dot), superimposed atop the the initial joint distribution of the uncorrelated Gaussian case. **(B)** Using the discrete model, we fix the mean susceptibility *ε* = 1 and show that varying the transmissibility of the index cases chosen in A can shift the timing of epidemic onsets and peaks. **(C)** Using stochastic simulations with a randomly chosen index case, we show the probability of an outbreak of more than 50 infections occurring for the SIR model, and for models incorporating negative correlations (*ρ* = −0.60), no correlations (*ρ* = 0) and positive correlations (*ρ* = 0.60) between susceptibility and transmissibility. **(D)** from the same stochastic ensembles as in C, variation in the final outbreak size (given that outbreaks generate more than 50 infections) is shown, outliers are shown as jittered points. Asterisk’s (*) show the results from the corresponding deterministic simulations from Figure 4. Parameters: transmission rate is *β* = 0.2 and recovery rate is *γ* = 0.1. Stochastic simulations were initialized in a population of 10,000. Each stochastic ensemble consists of 1,000 trajectories.

However, the susceptibility (and transmissibility) of individuals in the first few chains of infection may be important. To examine this, we adapted our model to include individual transmission events – utilising stochastic Gillespie simulations (see Appendix E) for the baseline SIR, and the heterogeneous cases with *ρ* = −0.6, *ρ* = 0, *ρ* = 0.6 examined in Figure 4. In doing so, we show that incorporating heterogeneity can additionally alter the probability of an outbreak (here defined as more than 50 infections, see Figure 5C). While the SIR model and the uncorrelated (*ρ* = 0) model both have an initial reproduction number of ℛ_0_ = 2, they differ in the likelihood of an outbreak occurring. For the SIR model, the outbreak probability (given *m* initial infections) is expected as: 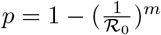 (Southall et al., 2023). With one initial index infection (*m* = 1) this supports an analytic outbreak probability of *p* = 0.5, in close agreement with the proportion of stochastic SIR model simulations in which an outbreak occurred (0.483); but further from the uncorrelated model (0.412). The correlated models also differ in the observed proportion of outbreaks relative to the expected outbreak probability; 0.339 vs *p* = 0.371 for the *ρ* = −0.6 case, and 0.474 vs. *p* = 0.5887 for the *ρ* = 0.6 condition. Regardless of correlation, the introduction of heterogeneity lowers the expected outbreak probability relative to the SIR baseline. Utilizing a stochastic framework also allows us to assess variability in epidemic trajectories (see Figure 5D) whose average final sizes for simulations that ran to epidemic burnout (i.e., simulations which ended due to susceptible depletion, rather than fizzling out) are in good agreement with the deterministic simulations. Histograms of final outbreak size and outbreak duration for all epidemic trajectories are shown in Figure S3. Together with Figure 4 we observe that within our framework, positive correlations between susceptibility and transmissibility lead to epidemics that are more likely to occur, and are faster (with shorter duration), stronger (higher incident infections and final outbreak size) and less variable (final outbreak size interquartile range = 0.0112, for *ρ* = 0.6), while negative correlation outbreaks are on average less likely to occur, have longer duration and lower, but more variable final size (final outbreak size interquartile range = 0.0239, for *ρ* = −0.6).

### 3.5 The effects of correlations on epidemic progress

Initial correlations between susceptibility and transmissibility of the population impact the strength (Figure 4) and potential (Figure 5) of outbreaks. However, correlations have consequences throughout an outbreak. Hence, we next explore how correlations between susceptibility and transmissibility impact epidemic trajectories. To do so, we vary the correlation coefficient, *ρ*, between *ρ* = −0.6 and *ρ* = 0.6 and match the exponential growth rate of incident infections by adjusting the transmission rate, *β*, to ensure an equivalent basic reproduction number (ℛ_0_ = 2) (see Figure 6A).We compare the epidemic dynamics during susceptible depletion using the progress variable, 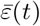, i.e., the mean susceptibility (Figure 6B). For 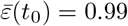, and 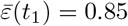, scenarios with positive (light blue), negative (dark blue), and no correlations (blue) reach this susceptibility level at a similar rate; but trajectories diverge as these scenarios move toward 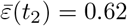. For negative correlations, we find the effective transmission rate increases over time, whereas for positive correlations, the effective transmission rate decreases over time (Figure 6C). The justification for this can be seen from the temporal evolution of the bivariate susceptibility-transmissibility distributions (shown by the changes in *f*_*S*_ with respect to 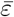 in Figure 6D). As the epidemics progress, the most susceptible individuals are more likely to be infected causing a reduction in mean susceptibility and effectively shifting the underlying distributions. The most salient effect here is the reduction of mean susceptibility over time as the most susceptible individuals become infected and then removed. With positive correlations, the most susceptible individuals are also the most transmissible; thus, as the most transmissible individuals are sculpted into the epidemic, the mean effective transmission rate of the remaining population declines. On the other hand, under negative correlations the effective transmission rates increase over time, as the least transmissible individuals are on average sculpted into the epidemic sooner such that the remaining population has higher average transmissibility. In Figure S4 and Figure S5 we examine these effects on population distributions by showing how the marginal susceptibility and transmissibility distributions in these particular models evolve with the mean susceptibility over time: At mean susceptibility, 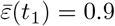, the susceptible population is depleted to about 80% of the population (Figure S4D). The marginal distributions for susceptibility are similar across *ρ* = − 0.6 to *ρ* = 0.6, with small deviations which we attribute to differences in truncation associated with the different correlation coefficients (Figure S4E). We find the marginal distributions for transmissibility differ at 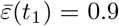 with positive correlation the transmissibility distribution is shifted to the right, whereas with negative correlations the transmissibility distribution is shifted to the left relative to the transmissibility distribution without correlations (Figure S4F). Since more susceptible individuals are infected earlier during the epidemic with positive correlations, the more susceptible individuals are also more transmissible, leading to increases in the initial speed and strength of the outbreak. With negative correlations, individuals who are more susceptible are less transmissible, leading to decreases in the initial speed and strength. We also further examined the susceptibility and transmissibility marginal distributions over time, comparing across simulations using the epidemic progress variable at values: 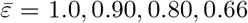 (Figure S5A, middle). For different correlation coefficients, the mean susceptibility, 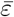, decreases at different rates due to differences in the dynamics of the effective transmission rates (Figure S5A, bottom). We show the susceptibility (Figure S5B) and transmissibility distributions (Figure S5C) over the epidemic progress variable (panels going down): Without correlations (*ρ* = 0), the effective transmission rate remains constant (Figure S5A). For positive correlations (*ρ* = 0.6), the effective transmission rate decreases over time, whereas for negative correlations (*ρ* = − 0.6), the effective transmission rate increases over time. Hence, in either case, the transmissibility distributions tend toward the mean transmissibility of 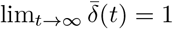 (Figure S5C), despite opposite tendencies in the effective transmission rate (Figure S5A, bottom panel). We note these differences in the temporal evolution of the marginal transmissibility distributions are offset by the differences in average transmission rate, *β* (which is 50% larger when *ρ* = −0.6 than when *ρ* = 0.6) in order to have the same ℛ_0_. However, the underlying differences in correlations cause the differences in effective transmission rate to evolve in opposite directions over time, which contribute to larger final outbreak sizes under negative correlations (Figure S4A).

**Figure 6.**
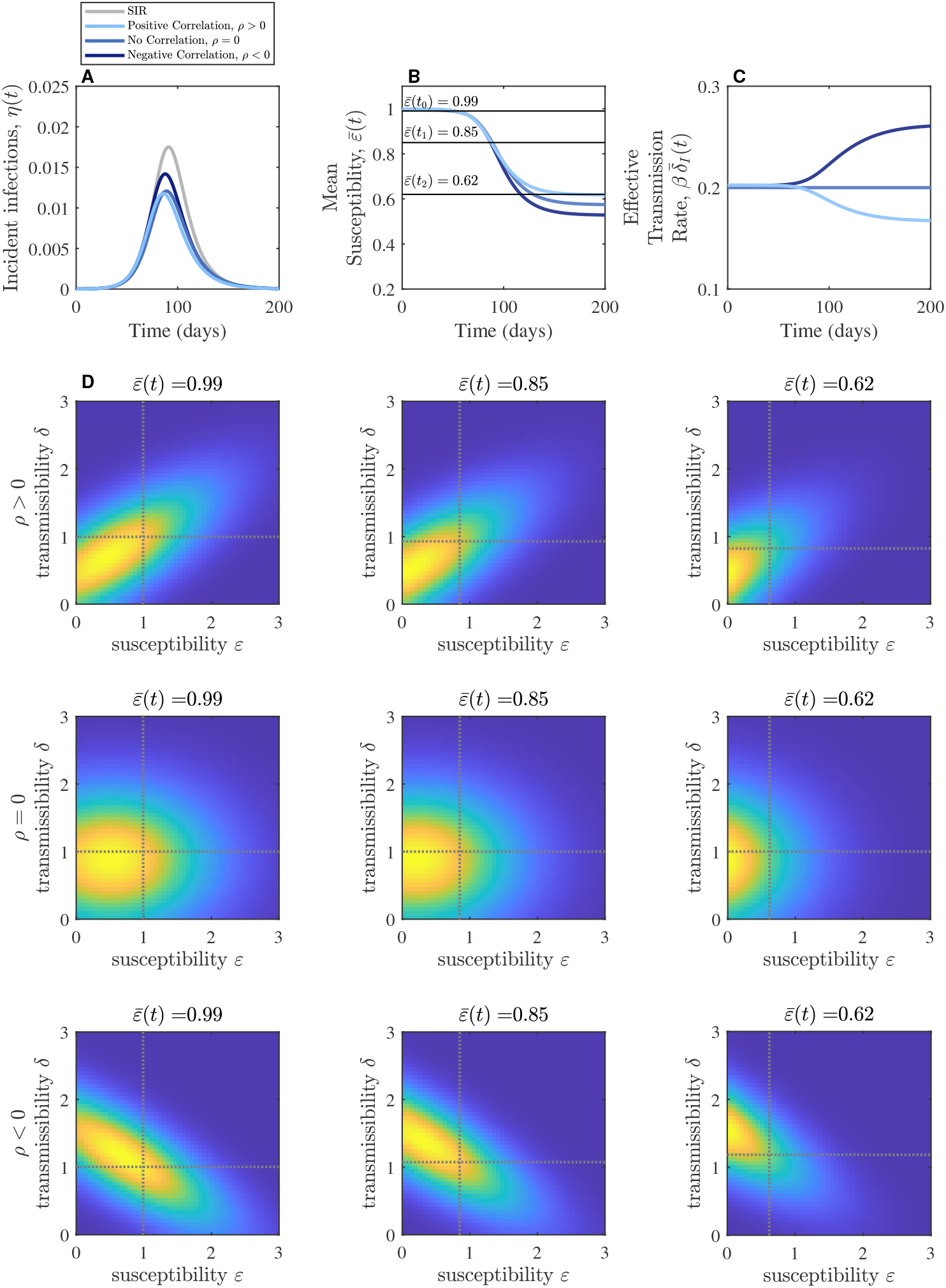
Temporal evolution of correlated susceptibility-transmissibility distributions under fixed ℛ_0_ = 2. **(A)** Incident infections. **(B)** Mean susceptibility decreases over time as the susceptible population is depleted. **(C)** Without correlations, the effective transmission rate remains constant. Positive correlations cause the transmissibility to decrease over time, whereas negative correlations cause transmissibility to increase over time. **(D)** bivariate distributions of *f*_*S*_ over time for *ρ* = 0.6 (top row), *ρ* = 0 (middle row), *ρ* =− 0.6 (bottom row) at positions marked by 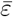 in B (columns: left 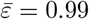, middle 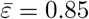, right 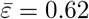). Dashed lines denote average susceptibility and transmissibility.

Overall, simulations of the full PDE model (Equation 11) agree with the qualitative analysis (see Supplementary Information). In particular, correlations modify the speed of susceptible depletion such that the epidemic slows down with positive correlations and speeds up with negative correlations between susceptibility and transmissibility. Consistent with results on heterogeneity in susceptibility, the final outbreak sizes are all less than in the conventional SIR model (see Figures 1-6). In this example, the uncorrelated case leads to about 60% of the initial susceptible population becoming infected, whereas the SIR model leads to about 80% of the susceptible population becoming infected (Figure S6A).

## 4 Discussion

We developed an epidemic model framework incorporating population-level covariation in both individual susceptibility and transmissibility. Our work investigates how susceptibility and transmissibility distributions are “sculpted” over the course of an epidemic, and how correlated variation may affect population-level dynamical outcomes. Consistent with prior findings (Rose et al., 2021; Gomes et al., 2022) initial gamma and Gaussian distributions are eigendistributions of the force of infection such that dynamics given uncorrelated susceptibility and transmissibility are equivalent to those in which average transmissibility is fixed and only susceptibility varies. Moving to exploring covariation, we identified a relation between ℛ_0_ and the correlation of initial potential transmissibility and susceptibility. Holding transmission and recovery rates constant, we found that when susceptibility and transmissibility are correlated (anticorrelated), then ℛ_0_ increases (decreases), epidemics initially grow faster (slower) and are more (less) likely to become outbreaks, and infect more (fewer) individuals. However, if instead ℛ_0_ is kept constant, we find models with covariation share the same initial epidemic speed, but differ in outcome. In order to keep ℛ_0_ fixed, and introducing negative correlations between susceptibility and transmissibility necessitates increasing the average transmission rate *β* (or decreasing the average recovery rate *γ*), leading to larger epidemics and additionally leading to an increase in the effective transmission rate over time, as individuals who are both highly susceptible and less transmissible are infected earlier on, leaving behind a less susceptible, but more transmissible population. Congruently and counterintuitively, when ℛ_0_ is fixed with positive correlations between susceptibility and transmissibility, epidemics are smaller than for the uncorrelated case and the effective transmission rate decreases over time.

This approach comes with caveats, insofar as we focus on inherent differences in individual susceptibility and transmissibility in a well-mixed population without vital dynamics and without the potential for reinfections. Going beyond inherent differences, recent work has highlighted that individual-level susceptibility and transmissibility can be associated with human behavior via risk-perceptive decision making (Salomon et al., 2021; Stolerman et al., 2023). Importantly, coupling informed human behaviour with disease dynamics can lead to conditions where dynamic changes in susceptibility (via changing behaviors) can explain epidemic peaks, oscillations, and shoulder behaviors (Weitz et al., 2020; Berestycki et al., 2023). The current model does not allow individual susceptibility or transmissibility to change in time, unlike Weitz et al., 2020 (while neglecting heterogeneity) and Berestycki et al., 2023 (while neglecting variability in transmissibility). Incorporating reinfection and vital dynamics might also enrich the observed dynamics and could allow one to probe differences in heritability of epidemiologically relevant life-history traits. While epidemic burnout is expected in well-mixed SIR models, even with vital dynamics (Parsons et al., 2024), population contact structure is also a highly relevant driver of disease dynamics (Keeling & Eames, 2005; Bansal et al., 2007; Funk et al., 2010; Prem et al., 2021). Our framework does not explicitly represent contact heterogeneity, however, variation in contact rates could be incorporated by assuming a positive correlation between susceptibility and transmissibility – with those who interact the most having the greatest potential to both catch and to transmit an infection. The degree to which contact rate variation factors as the strongest determinant in structuring the underlying joint distribution in susceptibility and transmissibility remains elusive and may differ between diseases and across contexts – however, our expectation is that for a naïve and fully susceptible population there are likely positive correlations between susceptibility and transmissibility. Future extensions might consider additional dynamical effects caused by incorporating additional parameter covariation with recovery rates, heterogeneity in vaccination (Saad-Roy et al., 2024), social determinants of health (Manna et al., 2024; Surasinghe et al., 2024), or with explicit population contact structure. Additionally, further investigation of how susceptibility and transmissibility distributions connect to other distributions of interest, such as the secondary attack rate (Anderson et al., 2023), is warranted.

There are also important questions to be addressed related to parameter inference and outbreak control. As we and others have shown, incorporating individual-level variation provides departures from baseline SIR dynamics (Dushoff, 1999; Novozhilov, 2008; Novozhilov, 2012; Karev & Novozhilov, 2019; Britton et al., 2020; Rose et al., 2021; Gomes et al., 2022; Anderson et al., 2023). In early outbreaks ℛ_0_ is one of the first parameters epidemiologists attempt to infer, yet our framing suggests ℛ_0_ might be entangled with covariation in susceptibility and transmissibility. For an identified value of ℛ_0_, we might expect different epidemic trajectories depending on the degree of covariation in the population. On the other hand, if ℛ_0_ is identified via average estimations of transmission and recovery rates, the degree of co-variation in the population may lead to mischaracterization of ℛ_0_. Beyond covariation in population-level susceptibility and transmissibility distributions, heterogeneities in population contact structure also factor into structuring transmission chains, which is not captured in our current models that assume well-mixed populations. Indeed, in network contexts ℛ_0_ is dependent on both the mean and variance of the degree distribution, as well as the correlation between vertex in- and out- degrees (Allard et al., 2023). Utilizing new inference approaches and data types will be required to identify the degree of covariation between relevant disease parameters e.g., (Kuylen et al., 2022; Anderson et al., 2023; Quilty et al., 2024; Tran-Kiem & Bedford, 2024; Tuschhoff & Kennedy, 2024). Beyond inference of ℛ_0_ as an early indicator of implementing control measures, there may be additional ramifications if susceptibility or transmissibility covary with infection severity. With a public health goal of minimizing severe outcomes across populations, then if severity is correlated with susceptibility and/or anti-correlated with transmissibility then stronger control measures may be required.

In closing, our work shows how covariation in heterogeneous levels of susceptibiliy and transmissibility scales up to population-level epidemics. Identifying dynamical hallmarks of covariation, and quantifying how multi-dimensional (dynamical) covariation drives population dynamics offer important future avenues to explore. In particular, given the relevance of heterogeneity to the liftoff and potential control of epidemics, we anticipate that decomposing the mechanistic basis of (co)variation of susceptibility and transmissibility will have both fundamental and applied relevance.

## Data Availability

MATLAB (version 2023b and 2024a) code for the analysis performed in this manuscript is available at https://github.com/Jeremy-D-Harris/SIR_heterogeneity_project and is archived on Zenodo, doi:10.5281/zenodo.13891898.

https://doi.org/10.5281/zenodo.13891898

## Authorship contribution statement

**Jeremy D. Harris:** Conceptualization, Formal analysis, Investigation, Software, Supervision, Visualization, Writing – original draft, Writing – review & editing. **Esther Gallmeier:** Conceptualization, Formal analysis, Investigation, Software, Visualization, Writing – review & editing. **Jonathan Dushoff:** Writing – review & editing. **Stephen J. Beckett:** Conceptualization, Investigation, Software, Validation, Visualization, Writing – original draft, Writing – review & editing. **Joshua S. Weitz:** Conceptualization, Funding acquisition, Project administration, Writing – review & editing.

## Acknowledgements

We thank Tapan Goel for constructive feedback and Raunak Dey for code review. This work was supported by grants from the National Science Foundation (no. 2200269) to JSW, the Simons Foundation (no. 930382) to JSW, and support from the Charities in Aid Foundation, the Marier Cunningham Foundation, and the Chaires Blaise Pascal program of the Île-de-France region (Blaise Pascal Institute Chair of Excellence awarded to JSW).

## Code availability

MATLAB (version 2023b and 2024a) code for the analysis performed in this manuscript is available at https://github.com/Jeremy-D-Harris/SIR heterogeneity project and is archived on Zenodo (Harris et al., 2024).

## A Derivation of susceptibility and transmissibility from discrete model variables

First, we write the susceptible (*S*) infected (*I*), and recovered (*R*) populations in terms of discrete model variables: *S*_*i,j*_, *I*_*i,j*_, and *R*_*i,j*_, where *i* and *j* are indices for the discrete susceptibility value, *ε*_*i*_, and transmissibility value, *δ*_*j*_. Then the force of infection for the susceptible population with susceptibility level *ε*_*i*_ is

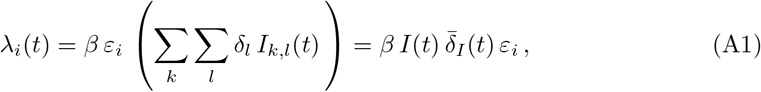

where *I*(*t*) is the total infected population and 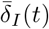 is the mean of the effective transmissibility distribution given by

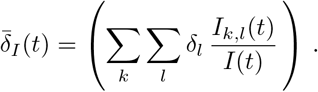

We can write the discrete model equations as

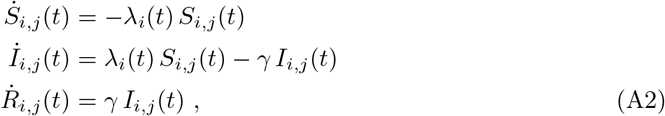

where *γ* is the recovery rate for infected individuals. This discrete model forms the basis of the continuous model equations given in Equation 11 with the connection between discrete and continuous model variables given as follows: *S*_*i,j*_ = *S*(*t, ε*_*i*_, *δ*_*j*_), *I*_*i,j*_ = *I*(*t, ε*_*i*_, *δ*_*j*_), and *R*_*i,j*_ = *R*(*t, ε*_*i*_, *δ*_*j*_). We can calculate the the total incidence:

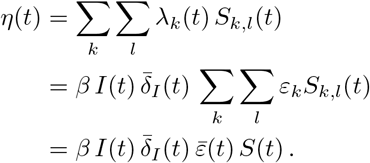

Next, we derive differential equations for 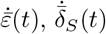 and 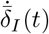 found in Equation 13 in terms of discrete model variables. For 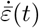, we examine the time derivative of total susceptibility,

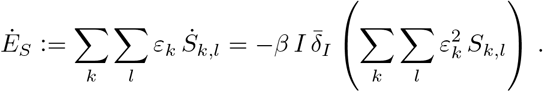

Note that 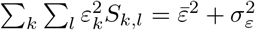. On the other hand, 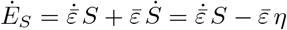. Hence,

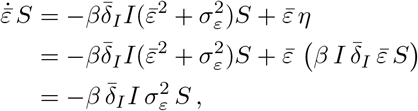

which leads to

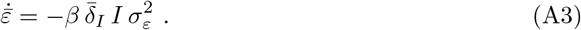

For 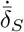, we examine the time derivative of the total potential transmissibility in the susceptible population,

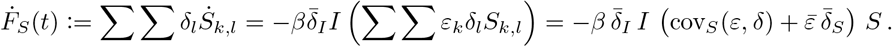

Here, cov_*S*_(*ε, δ*) is the covariance between susceptibility and transmissibility in the susceptible population. On the other hand, 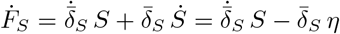. Hence,

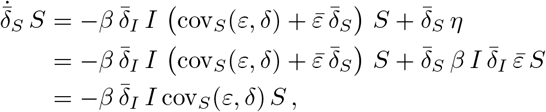

so that

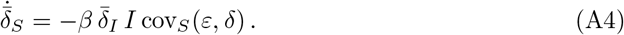

For 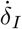, we examine the time derivative the total effective transmissibility in the infected population,

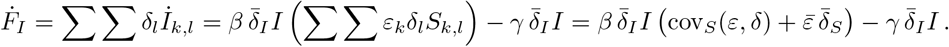

On the other hand, 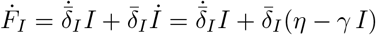. Equating sides and simplifying, we obtain

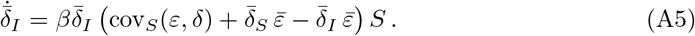

## B Derivations associated with Equation 19 and Equation 20

### B.1 Derivation of Equation 19

We derive the partial differential equation that describes the evolution of the susceptible sub-population density *f*_*S*_(*t, ε, δ*) with intrinsic susceptibility *ε* and intrinsic transmissibility *δ*. Rearranging the definition of *f*_*S*_(*t, ε, δ*) from Equation 1 and taking the partial derivative with respect to time, we obtain

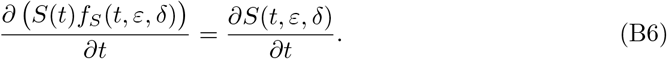

Expansion of the left-hand side through the product rule and the use of Equation 11 give us

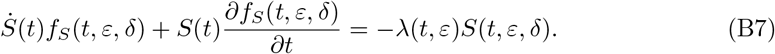

Recalling the definition of 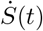 described in Equation 13, and expanding *λ* from Equation 10, we obtain

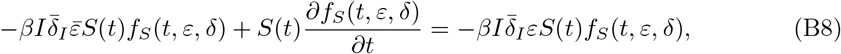

which after rearrangement yields the partial differential equation in Equation 19.

### B.2 Gamma and Gaussian distributions satisfy Equation 20

We check that gamma distributions with fixed shape parameter and Gaussian distributions with constant variance satisfy Equation 20. Consider setting the marginal susceptibility distribution as a gamma distribution:

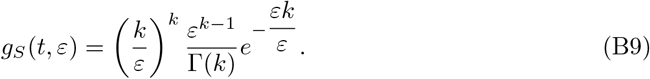

Fixing the susceptibility *ε* and the shape parameter *k*, we first take the partial derivative with respect to time *t* and apply the product rule. Subsequently, using Equation 11 (i.e., 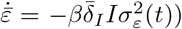 and the fact that 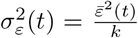 for gamma distributions, this expression simplifies and we recover Equation 20:

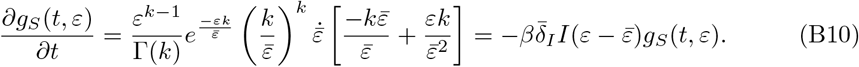

Similarly, consider setting the marginal susceptibility distribution as a Gaussian distribution:

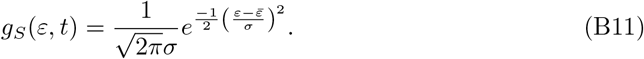

Fixing the susceptibility *ε* and the standard deviation *σ* (due to constant variance) and taking the partial derivative with respect to time *t*, and again recalling 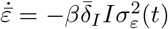 (see Equation 11), we show Gaussian distributions also satisfy Equation 20:

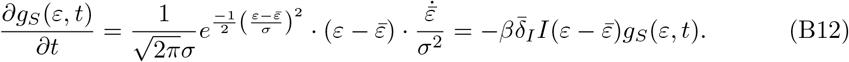

## C Initial joint distributions of susceptibility and transmissibility

To incorporate variation in both susceptibility and transmissibility we use bivariate distributions to initialize our epidemiological models. We do so by first creating the initial joint bivariate distribution in S, using a built-in function from the Statistics and Machine Learning Toolbox (MATLAB version 2023b and 2024a) for the specific probability distributions used. Without correlations between susceptibility and transmissibility, the initial joint distributions in *f*_*I*_ and *f*_*R*_ are set equal to that in *f*_*S*_. When correlations are introduced, the initial conditions of *f*_*I*_ and *f*_*R*_ are approximated via the attracting eigendistribution during the exponential growth phase of the epidemic from a preliminary simulation. In this work, we consider three types of bivariate distributions: gamma, truncated Gaussian, and negative binomial.

We first consider independent gamma distributions for the initial joint distribution for susceptibility (*ε*) and transmissibility (*δ*). However, these independent gamma distributions do not allow for covariation in *ε* and *δ*. Hence, we consider the bivariate Gaussian distribution with 2*×*2 covariance matrix, allowing us to compare the effects of increasing covariation on epidemic dynamics. We increase the initial variance in susceptibility (*σ*_*ε*_(0)) and transmissibility (*σ*_*δ*_(0)) to better see the effects of covariation. In doing so, the bivariate Gaussian distributions are truncated, because their support lies on the whole real plane ℝ ^2^. We ensure that mean values are set with 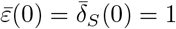 and adjust variances to match the specified correlation coefficient. In practice, truncation of distributions with high variance in susceptibility and/or transmissibility can be hard to match with large (anti)correlations. In our work, we restrict analysis to correlations between −0.6 and 0.6.

## D Model parameters

For all figures, except for Figure 4 which shows the reproduction number as a function of correlation coefficient, we set ℛ _0_ =2.0, a value representative of several respiratory viruses such as flu and SARS. We assume that the average time to recovery is 10 days and is exponentially distributed so that the recovery rate is given by *γ* = 1*/*10. Hence, we set *β* = 0.2, with the exception of Figure 6 (and related Supplemental figures) where *β* is adjusted to compensate for the effects of correlations (between susceptibility (*ε*) and transmissibility (*δ*)) and therefore, match the effective exponential growth rate of the epidemics across simulations. The model parameters (descriptions, values, and ranges, thereof) are shown in Table D1.

**Table D1.** Epidemiological model parameters. Epidemic parameters and distribution parameters explored in models with individual traits of susceptibility (*ε*) and transmissibility (*δ*), with 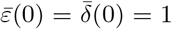. Baseline values refer to those used in the reference SIR model. Ranges indicate that these parameters vary based on initial conditions which depend on the correlation coefficient. Specific parameter values are noted in Figures.

| Parameter | Baseline value | Values explored | Description |
| --- | --- | --- | --- |
| $\mathcal{R}_0$ | 2.0 | 1.5 to 2.5 | Basic reproduction number |
| $\gamma$ | 1/10 day<sup>-1</sup> | 1/10 day<sup>-1</sup> | Recovery rate |
| $\beta$ | 0.2 | 0.165 to 0.254 day<sup>-1</sup> | Transmission rate of infections |
| $\sigma_\varepsilon^2(0)$ | 0 | 0.15 to 0.50 | Initial variance in susceptibility |
| $\sigma_\delta^2(0)$ | 0 | 0.05 to 0.35 | Initial variance in potential transmissibility |
| $\rho$ | NA | -0.60 to 0.60 | Correlation between susceptibility and potential transmissibility |

## E Simulation methods

All simulations and analysis were performed using MATLAB (version 2023b and 2024a). All simulation code is available at https://github.com/Jeremy-D-Harris/SIR heterogeneity project and archived on Zenodo (Harris et al., 2024).

### E.1 Deterministic simulations

To approximate the continuous susceptibility and transmissibility model variables, we use discrete variables composed of 100 uniformly spaced values between 0 and 6, such that the initial joint distributions we consider are seeded onto a uniform mesh of size 100 *×* 100. In visualization of the initial joint distributions we show only the range ∈ ([0, 3], [0, 3]), which represents ≈ 90% of the population. In all cases, distributions are chosen such that the initial population average susceptibility 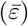 and potential transmissibility 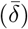 are equal to 1. Epidemic model simulations were numerically integrated using ode45 in MATLAB (Dormand & Prince, 1980; Shampine & Reichelt, 1997).

To implement initial conditions, we first create the initial joint distributions of *ε* and *δ* in the *S, I*, and *R* classes (see Appendix C). In all simulations except for in Figure 5, we let the total population be *N* = 1, as in Rose et al., 2021 and let the total population initial conditions be: *S* = *N, I* = 0, *R* = 0, with a small perturbation in the direction of the eigenvector of the SIR model. (Adjusting the perturbation magnitude translates the dynamics in time.) The initial conditions that are passed into the ode45 function are calculated from Equation 1 – Equation 3.

### E.2 Stochastic simulations

To analyze the outbreak potential of epidemics with different underlying susceptibility and transmissibility characteristics we utilize a stochastic simulation approach using the Gillespie algorithm (Gillespie, 1976, 1977). We initialize simulations with a population of 10,000 whose susceptibility and transmissibility values are seeded with probabilities taken from the joint probability distributions used in discrete model simulations to characterize a representative population with explicit individual-level variation. In each stochastic simulation run, one individual, chosen at random, is designated as the index infection. For each of the initial distributions we analyze (SIR, *ρ* = − 0.6, *ρ* = 0, *ρ* = 0.6) we run the stochastic simulation 1,000 times to obtain ensembles of epidemic trajectories; and denote a threshold of 50 infections to represent the occurrence an outbreak.

## Supplemental Material

From equations Equation 11 and Equation 13, the joint distribution in the infected population satisfies the partial differential equation:

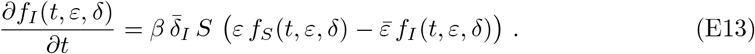

Integrating over *ε*, the effective transmissibility distribution satisfies the partial differential equation:

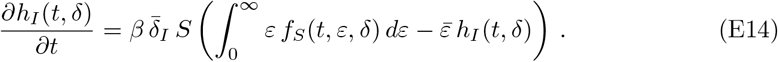

In the case of uncorrelated susceptibility and transmissibility values, 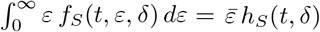, which means that the effective transmissibility distribution remains constant and equal to the potential transmissibility distribution in the susceptible population, if they are initially equal. The mean effective transmissibility remains constant, here equal to 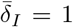 (Figure S5C; medium blue dot). For positive correlations, the mean effective transmissibility is greater than in the case without correlations (Figure S5C; light blue dot), whereas for negative correlations, the mean effective transmissibility is less than in the case without correlations (Figure S5C; dark blue dot). For positive (negative) correlations between susceptibility and transmissibility, initial incident infections are comprised of more (less) transmissible individuals. Thus, for positive correlations, 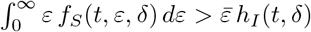 such that 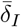 monotonically decreases toward 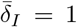. whereas for negative correlations, 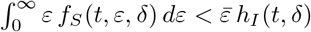 such that 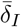 monotonically increases toward 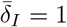.

## Supplemental Figures

**Figure S1.**
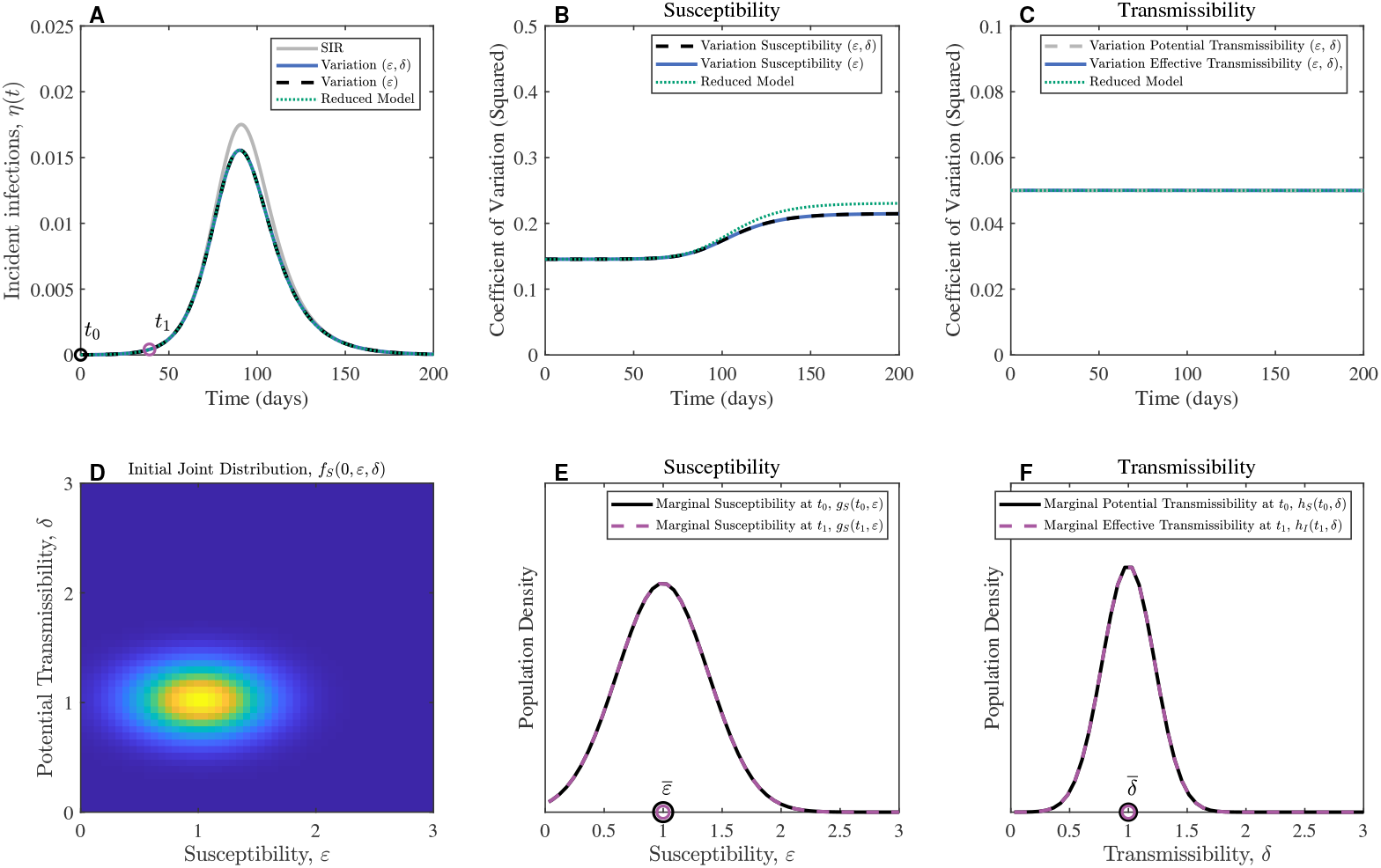
Uncorrelated (low variance) Gaussian distributions for susceptibility and transmissibility during exponential growth. Population dynamics with variation in susceptibility and transmissibility. **(A)** Incident infections. **(B)** Coefficient of Variation (squared) in susceptibility remains constant during exponential growth and increases over time as the susceptible population decreases. **(C)** When transmissibility and susceptibility are uncorrelated, Coefficient of Variation (squared) in transmissibility remains constant over time for Gaussian distributions. **(D)** Initial joint distributions (uncorrelated Gaussian distributions) of susceptibility values (*ε*) and potential transmissibility values (*δ*). **(E)** Susceptibility distributions remain constant during exponential growth, shown at two time points: *t*_0_ = 0 and *t*_1_ = 50 days. **(F)** Potential and effective transmissibility distributions at the time points: *t*_0_ = 0 and *t*_1_ = 50 days. The transmission rate is equal to *β* = 0.2, and the recovery rate is equal to *γ* = 1*/*10 such that the basic reproduction number is ℛ_0_ = 2.0. The variance values in the initial joint: 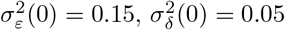. The reduced model refers to Equation 18 with 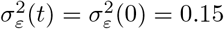.

**Figure S2.**
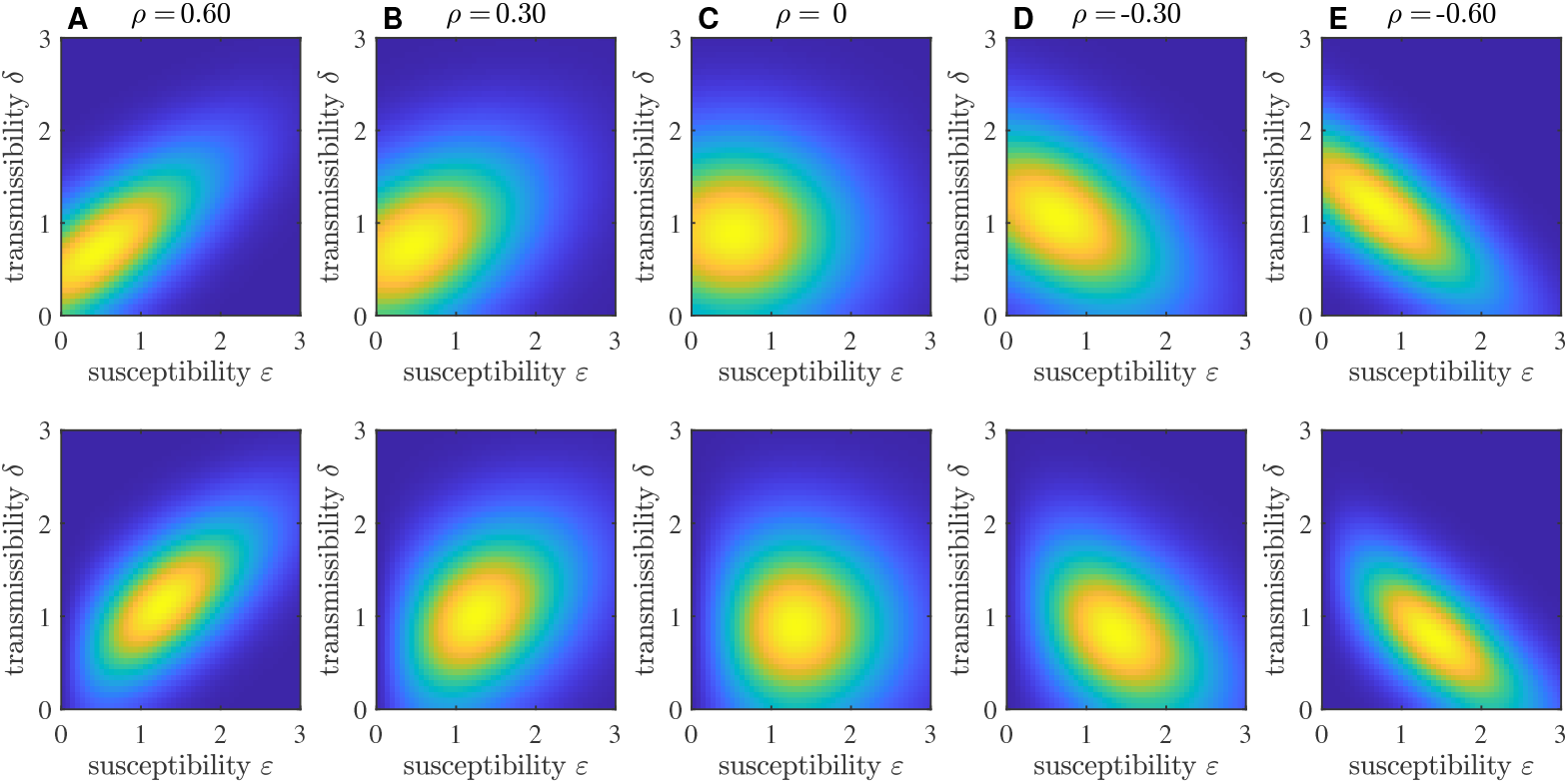
Initial Gaussian Joint Distributions in *S* (top) and *I* (bottom). Corresponding to simulations in Figure 4, where the joint distribution in *I* arises from the eigendistribution: (**A**) *ρ* = −0.6, (**B**) *ρ* = −0.3, (**C**) *ρ* = 0, (**D**) *ρ* = 0.3, (**E**) *ρ* = 0.6.

**Figure S3.**
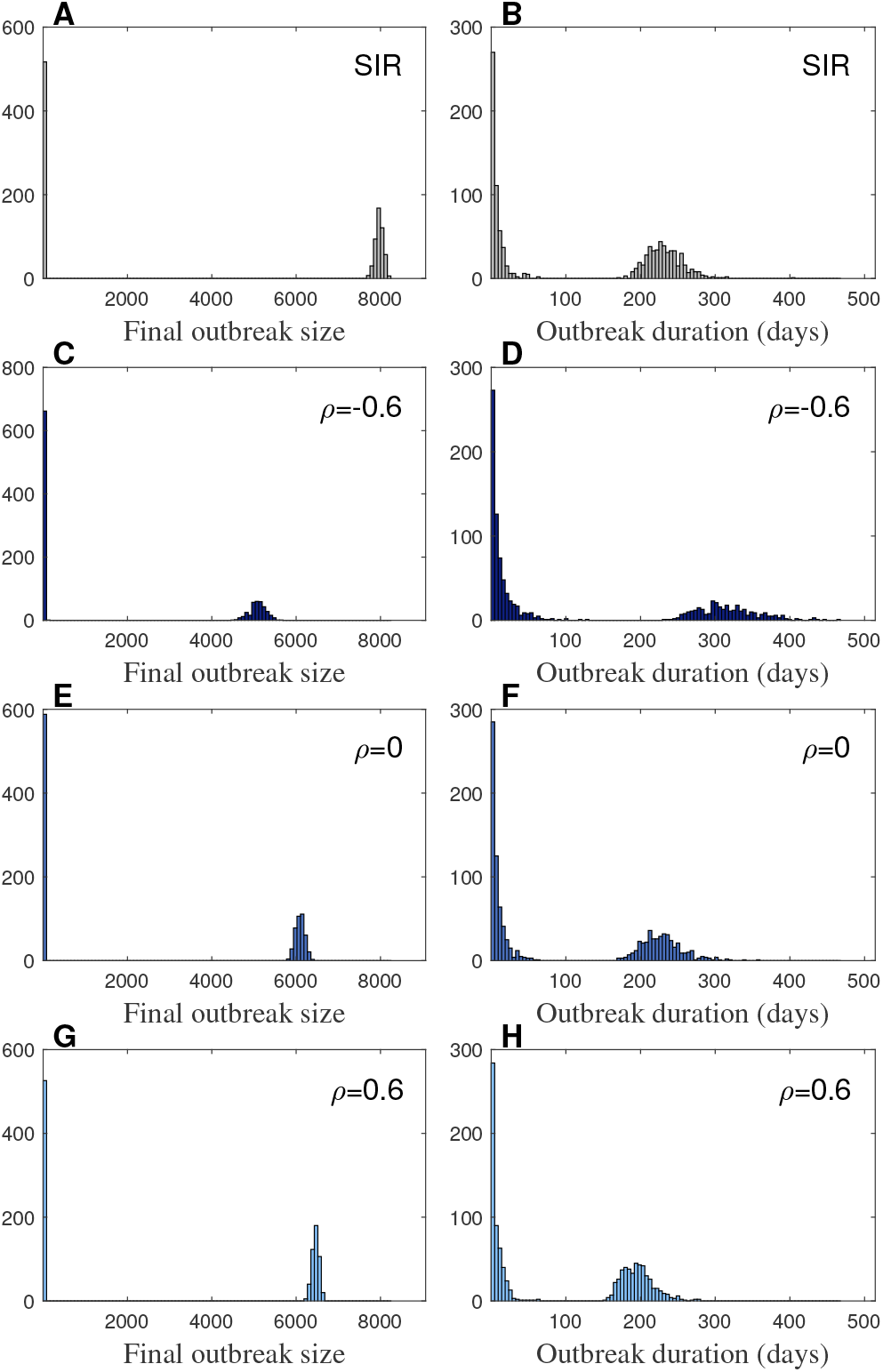
Stochastic variability in epidemic outbreak size and duration. Count histograms from the four ensembles of 1,000 simulations of epidemic trajectories shown in Figure 5C,D. Final outbreak size **(A)** and duration (days) **(B)** for the SIR model, where 51.7% of simulations generated ≤ 50 infections. Final outbreak size **(C)** and duration (days) **(D)** for the model with negative correlation between susceptibility and potential transmissibility (*ρ* = − 0.6), where 66.1% of simulations generated ≤ 50 infections. Final outbreak size **(E)** and duration (days) **(F)** for the model with no correlation between susceptibility and potential transmissibility (*ρ* = 0), where 58.8% of simulations generated ≤ 50 infections. Final outbreak size **(G)** and duration (days) **(H)** for the model with positive correlation between susceptibility and potential transmissibility (*ρ* = 0.6), where 52.6% of simulations generated ≤ 50 infections. Parameters: transmission rate is *β* = 0.2 and recovery rate is *γ* = 0.1. Stochastic simulations were initialized in a population of 10,000.

**Figure S4.**
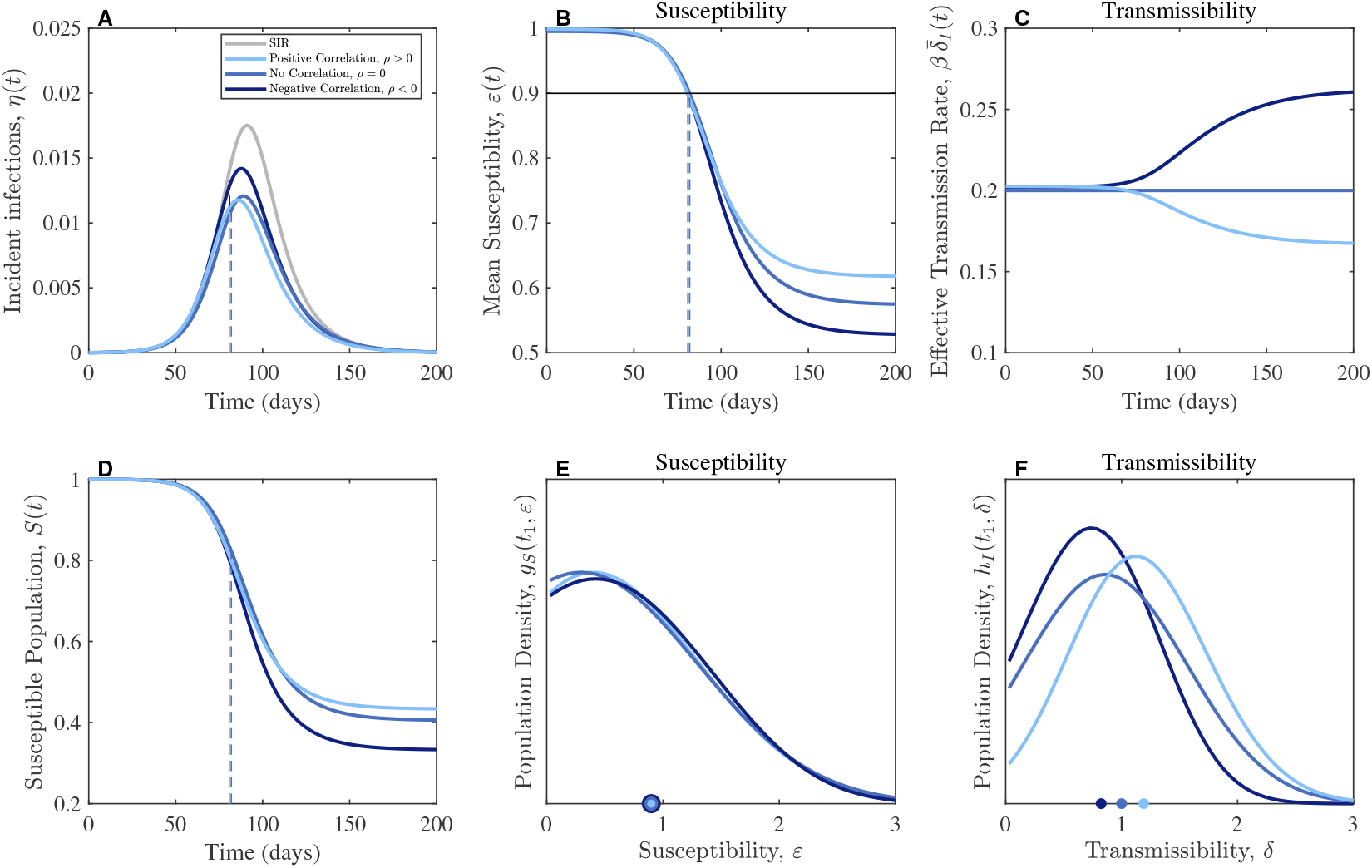
The effects of correlations between susceptibility and transmissibility when the exponential growth rate is matched. **(A)** Incident infections. **(B)** Mean susceptibility decreases over time as the susceptible population is depleted. **(C)** Without correlations, the effective transmission rate remains constant (medium blue). Positive correlations cause the transmissibility to decrease over time (light blue), whereas negative correlations cause transmissibility to increase over time (dark blue). **(D)** About 80% of the susceptible population is depleted. **(E)** Susceptibility distributions plotted at 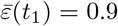 **(F)** Transmissibility distribution at time points corresponding to the progress variable, 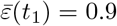 For positive (negative) correlations, the mean transmissibility is greater (less) than the mean transmissibility without correlations. Across all simulations, the recovery rate is *γ* = 1*/*10 and the basic reproduction number is ℛ _0_ = 2.0. The transmission rates vary: *β* = 0.254, 0.2, 0.165 for negative correlation (*ρ* = − 0.6), no correlation (*ρ* = 0), and positive correlation (*ρ* = 0.6), respectively. The initial variance values are: 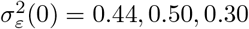 and 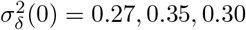.

**Figure S5.**
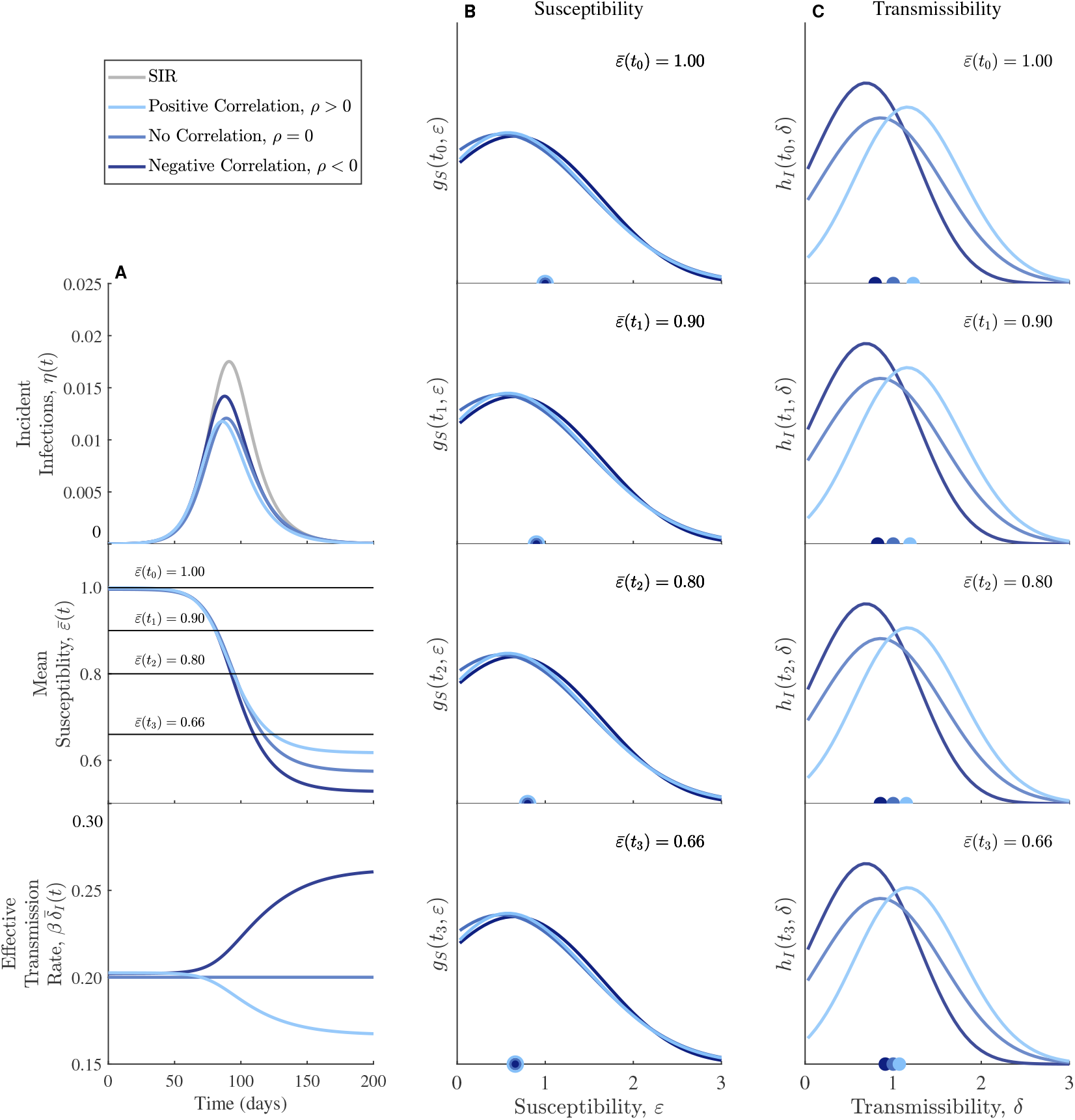
The effects of correlations on susceptibility and transmissibility distributions over time. **(A)** Population dynamics including incident infections (*η*), the mean susceptibility 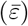 (both redrawn from Figure S4A,B), and the effective transmission rate 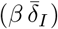. **(B)** Susceptibility distributions associated with four mean susceptibility values going down the rows: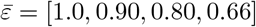,**(C)** Effective transmissibility distributions at the same four time points corresponding to the mean susceptibility values. Parameter values are the same as in Figure S4. Corresponding coefficients of variation (squared) are shown in Figure S6.

**Figure S6.**
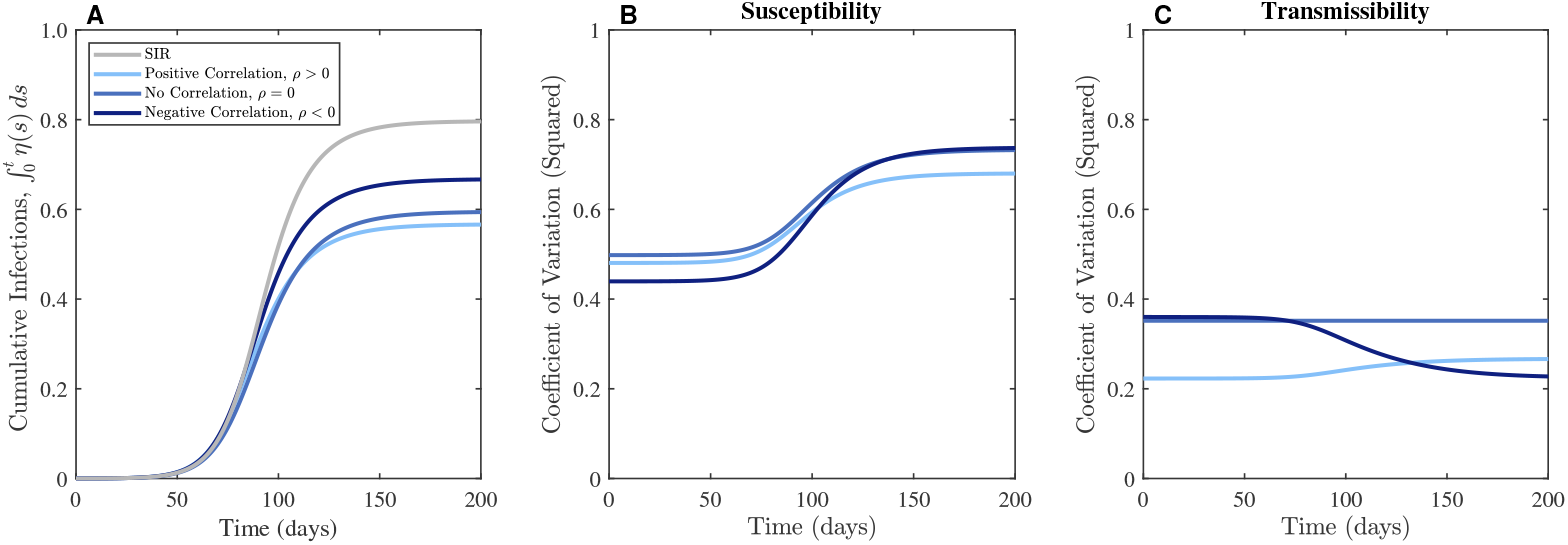
The effects of correlations on cumulative infections and the coefficients of variation. (**A**) Cumulative infections showing different final outbreak sizes for different correlations between susceptibility and transmissibility. Parameter values are the same as in Figure S4 and Figure S5. The coefficients of variation (squared) for susceptibility (**B**) and transmissibility (**C**).

## Notes

### Competing Interest Statement

The authors have declared no competing interest.

## References

Allard, A., Moore, C., Scarpino, S. V., Althouse, B. M., & Hébert-Dufresne, L. (2023). The role of directionality, heterogeneity, and correlations in epidemic risk and spread. SIAM Review, 65 (2), 471–492. 10.1137/20m1383811

Anderson, T. L., Nande, A., Merenstein, C., Raynor, B., Oommen, A., Kelly, B. J., Levy, M. Z., & Hill, A. L. (2023). Quantifying individual-level heterogeneity in infectiousness and susceptibility through household studies. Epidemics, 44, 100710. 10.1016/j.epidem.2023.100710

Ball, F. (1985). Deterministic and stochastic epidemics with several kinds of susceptibles. Advances in applied probability, 17 (1), 1–22. 10.2307/1427049

Bansal, S., Grenfell, B. T., & Meyers, L. A. (2007). When individual behaviour matters: Homogeneous and network models in epidemiology. Journal of The Royal Society Interface, 4 (16), 879–891. 10.1098/rsif.2007.110017/32

Berestycki, H., Desjardins, B., Weitz, J. S., & Oury, J.-M. (2023). Epidemic modeling with heterogeneity and social diffusion. Journal of Mathematical Biology, 86 (4), 60. 10.1007/s00285-022-01861-w

Britton, T., Ball, F., & Trapman, P. (2020). A mathematical model reveals the influence of population heterogeneity on herd immunity to SARS-CoV-2. Science, 369 (6505), 846–849. 10.1126/science.abc6810

Coutinho, F., Massad, E., Lopez, L., Burattini, M., Struchiner, C., & Azevedo-Neto, R. (1999). Modelling heterogeneities in individual frailties in epidemic models. Mathematical and computer modelling, 30 (1-2), 97–115. 10.1016/S0895-7177(99)00119-3

Davies, N. G., Klepac, P., Liu, Y., Prem, K., Jit, M., & Eggo, R. M. (2020). Age-dependent effects in the transmission and control of COVID-19 epidemics. Nature medicine, 26 (8), 1205–1211. 10.1038/s41591-020-0962-9

Dormand, J. R., & Prince, P. J. (1980). A family of embedded Runge-Kutta formulae. Journal of computational and applied mathematics, 6 (1), 19–26. 10.1016/0771-050X(80)90013-3

Dushoff, J. (1999). Host heterogeneity and disease endemicity: A moment-based approach. Theoretical Population Biology, 56 (3), 325–335. 10.1006/tpbi.1999.1428

Dushoff, J., & Levin, S. (1995). The effects of population heterogeneity on disease invasion. Mathematical Biosciences, 128 (1-2), 25–40. 10.1016/0025-5564(94)00065-8

Dwyer, G., Dushoff, J., Elkinton, J. S., & Levin, S. A. (2000). Pathogen-driven outbreaks in forest defoliators revisited: Building models from experimental data. The American Naturalist, 156 (2), 105–120. 10.1086/303379

Dwyer, G., Elkinton, J. S., & Buonaccorsi, J. P. (1997). Host heterogeneity in susceptibility and disease dynamics: Tests of a mathematical model. The American Naturalist, 150 (6), 685–707. 10.1086/286089

Eksin, C., Paarporn, K., & Weitz, J. S. (2019). Systematic biases in disease forecasting – the role of behavior change. Epidemics, 27, 96–105. 10.1016/j.epidem.2019.02.004

Endo, A., Abbott, S., Kucharski, A. J., & Funk, S. (2020). Estimating the overdispersion 54 in COVID-19 transmission using outbreak sizes outside China. Wellcome Open 54 Research, 5, 67. 10.12688/wellcomeopenres.15842.3

Frieden, T. R., & Lee, C. T. (2020). Identifying and interrupting superspreading events—implicati5o4 for control of severe acute respiratory syndrome coronavirus 2. Emerging Infectious 54 Diseases, 26 (6), 1059–1066. 10.3201/eid2606.200495

Funk, S., Salathé, M., & Jansen, V. A. (2010). Modelling the influence of human behaviour 55 on the spread of infectious diseases: A review. Journal of the Royal Society Interface, 55 7 (50), 1247–1256. 10.1098/rsif.2010.0142

Gart, J. J. (1972). The statistical analysis of chain-binomial epidemic models with several kinds of susceptibles. Biometrics, 28, 921–930. 10.2307/2528629

Gillespie, D. T. (1976). A general method for numerically simulating the stochastic time evolution of coupled chemical reactions. Journal of computational physics, 22 (4), 403–434. 10.1016/0021-9991(76)90041-3

Gillespie, D. T. (1977). Exact stochastic simulation of coupled chemical reactions. The Journal of Physical Chemistry, 81 (25), 2340–2361. 10.1021/j100540a008

Gomes, M. G. M., Ferreira, M. U., Corder, R. M., King, J. G., Souto-Maior, C., PenhaGonçalves, C., Gonçalves, G., Chikina, M., Pegden, W., & Aguas, R. (2022). Individual variation in susceptibility or exposure to SARS-CoV-2 lowers the herd immunity threshold. Journal of Theoretical Biology, 111063. 10.1016/j.jtbi.2022.111063

Goyal, A., Reeves, D. B., & Schiffer, J. T. (2022). Multi-scale modelling reveals that early super-spreader events are a likely contributor to novel variant predominance. Journal of The Royal Society Interface, 19 (189). 10.1098/rsif.2021.0811

Harris, J. D., Gallmeier, E., Dushoff, J., Beckett, S. J., & Weitz, J. S. (2024). Code for: “Infections are not alike: the effects of covariation between individual susceptibility and transmissibility on epidemic dynamics” [Zenodo, doi: 10.5281/zenodo.13891898]. 10.5281/zenodo.13891898

Karev, G. P., & Novozhilov, A. S. (2019). How trait distributions evolve in populations with parametric heterogeneity. Mathematical Biosciences, 315, 108235. 10.1016/j.mbs.2019.108235

Keeling, M. J., & Eames, K. T. (2005). Networks and epidemic models. Journal of the Royal Society Interface, 2 (4), 295–307. 10.1098/rsif.2005.0051

Kermack, W. O., & McKendrick, A. G. (1927). A contribution to the mathematical theory of epidemics. Proceedings of the Royal Society of London. Series A, Containing papers of a Mathematical and Physical Character, 115 (772), 700–721. 10.1098/rspa.1927.0118

Kuylen, E. J., Torneri, A., Willem, L., Libin, P. J., Abrams, S., Coletti, P., Franco, N., Verelst, F., Beutels, P., Liesenborgs, J., & Hens, N. (2022). Different forms of superspreading lead to different outcomes: Heterogeneity in infectiousness and contact behavior relevant for the case of SARS-CoV-2. PLoS Computational Biology, 18 (8), e1009980. 10.1371/journal.pcbi.1009980

Lloyd-Smith, J. O. (2007). Maximum likelihood estimation of the negative binomial dispersion parameter for highly overdispersed data, with applications to infectious diseases. PLoS ONE, 2 (2), e180. 10.1371/journal.pone.0000180

Lloyd-Smith, J. O., Schreiber, S. J., Kopp, P. E., & Getz, W. M. (2005). Superspreading and the effect of individual variation on disease emergence. Nature, 438 (7066), 355–359. 10.1038/nature04153

Lovell-Read, F. A., Shen, S., & Thompson, R. N. (2022). Estimating local outbreak risks and the effects of non-pharmaceutical interventions in age-structured populations: SARS-CoV-2 as a case study. Journal of Theoretical Biology, 535, 110983. 10.1016/j.jtbi.2021.110983

Manna, A., Koltai, J., & Karsai, M. (2024). Importance of social inequalities to contact patterns, vaccine uptake, and epidemic dynamics. Nature Communications, 15 (1). 10.1038/s41467-024-48332-y

Meehan, M. T., Hughes, A., Ragonnet, R. R., Adekunle, A. I., Trauer, J. M., Jayasundara, P., McBryde, E. S., & Henderson, A. S. (2023). Replicating superspreader dynamics with compartmental models. Scientific Reports, 13 (1), 15319. 10.1038/s41598-023-42567-3

Murayama, H., Pearson, C. A. B., Abbott, S., Miura, F., Jung, S.-m., Fearon, E., Funk, S., & Endo, A. (2023). Accumulation of immunity in heavy-tailed sexual contact networks shapes Mpox outbreak sizes. The Journal of Infectious Diseases, 229 (1), 59–63. 10.1093/infdis/jiad254

Novozhilov, A. S. (2012). Epidemiological models with parametric heterogeneity: Deterministic theory for closed populations. Mathematical Modelling of Natural Phenomena, 7 (3), 147–167. 10.1051/mmnp/20127310

Novozhilov, A. S. (2008). On the spread of epidemics in a closed heterogeneous population. Mathematical Biosciences, 215 (2), 177–185. 10.1016/j.mbs.2008.07.010

Parsons, T. L., Bolker, B. M., Dushoff, J., & Earn, D. J. (2024). The probability of epidemic burnout in the stochastic SIR model with vital dynamics. Proceedings of the National Academy of Sciences, 121 (5), e2313708120. 10.1073/pnas.2313708120

Prem, K., Zandvoort, K. v., Klepac, P., Eggo, R. M., Davies, N. G., Cook, A. R., & Jit, M. (2021). Projecting contact matrices in 177 geographical regions: An update and comparison with empirical data for the COVID-19 era. PLOS Computational Biology, 17 (7), e1009098. 10.1371/journal.pcbi.1009098

Quilty, B. J., Chapman, L. A., Munday, J. D., Wong, K. L., Gimma, A., Pickering, S., Neil, S. J., Galao, R., Edmunds, W. J., Jarvis, C. I., & Kucharski, A. J. (2024). Disentangling the drivers of heterogeneity in SARS-CoV-2 transmission from data on viral load and daily contact rates. bioRxiv. 10.1101/2024.08.15.24311977

Rose, C., Medford, A. J., Goldsmith, C. F., Vegge, T., Weitz, J. S., & Peterson, A. A. (2021). Heterogeneity in susceptibility dictates the order of epidemic models. Journal of Theoretical Biology, 528, 110839. 10.1016/j.jtbi.2021.110839

Saad-Roy, C. M., Morris, S. E., Boots, M., Baker, R. E., Lewis, B. L., Farrar, J., Marathe, M. V., Graham, A. L., Levin, S. A., Wagner, C. E., Metcalf, C. J. E., & Grenfell, B. T. (2024). Impact of waning immunity against SARS-CoV-2 severity exacerbated by vaccine hesitancy. PLOS Computational Biology, 20 (8), e1012211. 10.1371/journal.pcbi.1012211

Salomon, J. A., Reinhart, A., Bilinski, A., Chua, E. J., La Motte-Kerr, W., Rönn, M. M., Reitsma, M. B., Morris, K. A., LaRocca, S., Farag, T. H., Kreuter, F., Rosenfeld, R., & Tibshirani, R. J. (2021). The US COVID-19 trends and impact survey: Continuous real-time measurement of COVID-19 symptoms, risks, protective behaviors, testing, and vaccination. Proceedings of the National Academy of Sciences, 118 (51). 10.1073/pnas.2111454118

Shampine, L. F., & Reichelt, M. W. (1997). The MATLAB ODE suite. SIAM journal on scientific computing, 18 (1), 1–22. 10.1137/S1064827594276424

Sneppen, K., Nielsen, B. F., Taylor, R. J., & Simonsen, L. (2021). Overdispersion in COVID-19 increases the effectiveness of limiting nonrepetitive contacts for transmission control. Proceedings of the National Academy of Sciences, 118 (14). 10.1073/pnas.2016623118

Southall, E., Ogi-Gittins, Z., Kaye, A., Hart, W., Lovell-Read, F., & Thompson, R. (2023). A practical guide to mathematical methods for estimating infectious disease outbreak risks. Journal of Theoretical Biology, 562, 111417. 10.1016/j.jtbi.2023.111417

Stolerman, L. M., Clemente, L., Poirier, C., Parag, K. V., Majumder, A., Masyn, S., Resch, B., & Santillana, M. (2023). Using digital traces to build prospective and real-time county-level early warning systems to anticipate COVID-19 outbreaks in the United States. Science Advances, 9 (3). 10.1126/sciadv.abq0199

Surasinghe, S., Manivannan, S. N., Scarpino, S. V., Crawford, L., & Ogbunugafor, C. B. (2024). Structural causal influence (SCI) captures the forces of social inequality in models of disease dynamics. 10.48550/ARXIV.2409.09096

Tran-Kiem, C., & Bedford, T. (2024). Estimating the reproduction number and transmission heterogeneity from the size distribution of clusters of identical pathogen sequences. Proceedings of the National Academy of Sciences, 121 (15), e2305299121. 10.1073/pnas.2305299121

Tuschhoff, B. M., & Kennedy, D. A. (2024). Detecting and quantifying heterogeneity in susceptibility using contact tracing data. PLOS Computational Biology, 20 (7), e1012310. 10.1371/journal.pcbi.1012310

Weitz, J. S., Park, S. W., Eksin, C., & Dushoff, J. (2020). Awareness-driven behavior changes can shift the shape of epidemics away from peaks and toward plateaus, shoulders, and oscillations. Proceedings of the National Academy of Sciences, 117 (51), 32764–32771. 10.1073/pnas.200991111720/32

Wong, F., & Collins, J. J. (2020). Evidence that coronavirus superspreading is fat-tailed. Proceedings of the National Academy of Sciences, 117 (47), 29416–29418. 10.1073/pnas.2018490117

Zhang, J., Litvinova, M., Liang, Y., Wang, Y., Wang, W., Zhao, S., Wu, Q., Merler, S., Viboud, C., Vespignani, A., et al. (2020). Changes in contact patterns shape the dynamics of the COVID-19 outbreak in China. Science, 368 (6498), 1481–1486. 10.1126/science.abb8001

